# Association of bile acids, amino acids, and glycerophospholipid metabolites with food-allergic outcomes in children on peanut oral immunotherapy

**DOI:** 10.1101/2025.07.01.25330651

**Authors:** Jennifer N. Styles, Justin K. Wang, Ananya Raman, Michael Lu, Jessica A. Lasky-Su, Brian P. Vickery, Wesley A. Burks, Tim Moran, Mike Kulis, Wayne Shreffler, Edwin H. Kim, Yamini V. Virkud

**Affiliations:** University of North Carolina at Chapel Hill, Department of Pediatrics, Chapel Hill, NC; Massachusetts General Hospital for Children, Food Allergy Center. Department of Pediatrics, Massachusetts General Hospital, Boston, MA; Harvard Medical School, Boston, MA; Broad Institute, Cambridge, MA; Channing Division of Network Medicine, Brigham & Women’s Hospital, Boston, MA; Emory University and Children’s Healthcare of Atlanta, GA; University of North Carolina School of Medicine and UNC Health

## Abstract

**Background:** Food allergen immunotherapy can induce remission of food allergies in certain individuals, but the mechanisms underlying this remission are largely unknown. Prior work has identified differences in immunomodulatory metabolites between older children who develop remission versus non-remission on oral immunotherapy (OIT). Here we aim to characterize metabolomic changes during OIT in young children to and validate patterns of immunomodulatory metabolites previously observed.

**Methods:** Untargeted plasma metabolomic profiling was performed on samples from the DEVIL peanut OIT trial (n=41, ages 9-36 months). Remission status was determined by oral food challenges conducted at the end-of-therapy and the end of a 1-month avoidance period. Linear and logistic regression models were used to detect differences in individual metabolites over time on OIT and by remission status. Pathway analyses were used to determine enrichment of chemical subclasses and biological pathways. These pathways were then compared to prior findings generated from similar profiling performed from the PNOIT peanut OIT trial (n=20, ages 7-13).

**Results:** During OIT, glycerophospholipid metabolites (q=3.8×10^-5^) increased over time and most amino acid metabolites (q=6.1×10^-45^) decreased over time. Participants who went on to develop remission, had higher levels of amino acids (q=4.3×10^-8^) and bile acids (q=0.00014), whereas children who developed non-remission had higher levels of glycerophospholipids (q=4.3×10^-10^). Comparison of these findings with our second cohort of OIT in older children, showed replication of the enrichment of glycerophosphocholines (q=1.0×10^-13^) and amino acids (q=2.1×10^-5^) among metabolites that changed over time on OIT and replication of glycerophospholipids(q=5.7×10^-16^), amino acids (*PNOIT* q=7.2×10^-7^), and bile acids (*PNOIT* q=3.8×10^-8^) among metabolites that varied by remission status.

**Conclusions:** Metabolomic profiles on OIT differed both over time on OIT and by OIT remission status in young children. Between two independent OIT cohorts of different ages we observed replication of significantly enriched chemical subclasses of glycerophospholipids, amino acids, and bile acids. Given the potentially immunomodulatory roles of some of these metabolites, our results suggest that glycerophospholipids, amino acids, and bile acids may be involved in the mechanisms of remission on OIT.

## Introduction

Therapies to treat and cure food allergies are needed but are currently very limited.^1-5^ One form of allergen-specific immunotherapy, oral immunotherapy (OIT) was approved by the FDA in 2020 for peanut allergy.^6^ OIT consists of administering gradually increasing doses of peanut protein until reaching a maintenance dose designed to increase the allergen threshold of most patients.^7, 8^ However, while some people are able to sustain this increased threshold at the end of allergen-specific immunotherapy (‘remission’), for others the allergy returns soon after stopping treatment (‘transient desensitization’).^9–11^ These disease-modifying effects of OIT on allergen thresholds support the use of OIT trials to study how immune tolerance could be altered as a result of desensitization or remission.^9,11–14^

Several studies have shown higher rates of remission with younger ages, supporting the importance of studying the pathophysiology of remission in infants and toddlers.^9, 11, 15^ The Determining the Efficacy and Value of Immunotherapy on the Likelihood of Peanut Tolerance (*DEVIL*) trial was the first study of OIT in toddlers (9-36 months at enrollment). This trial demonstrated high rates of desensitization and 1-month remission (termed ‘sustained unresponsiveness’) that exceeded those reported for OIT in older children, without sacrificing safety.^11, 16^

One approach to study the mechanistic differences underlying desensitization and remission includes metabolomics, the study of small molecule ‘metabolites’ and specifically bioactive metabolites. Metabolomics allows us to look at the products of genetics, environmental exposures, diet, and other physiological and biological processes and how they are altered by OIT.^17^ These metabolites can also have active roles in altering these biological processes, including the immune system.^18, 19^ Global metabolomics, which identifies a broad range of detectable circulating metabolites, allows for the analysis of overarching patterns of chemical subclasses and biological pathways as well as individual metabolite trends.^17^

We and others have studied metabolomics in food allergy, and have detected differences in bile acids, amino acids, eicosanoids, plasmalogens, histidine metabolites and other chemical subclasses.^13, 20–23^ Specifically, in the first examination of plasma metabolomics profiles during OIT, we found that bile acids, eicosanoids, histidine metabolites, and glycerophospholipids were altered between older children (7-13years old) who developed remission (or sustained unresponsiveness) compared to transient desensitization.^13^ Many of these metabolites have immunomodulatory roles; for example bile acids regulate a variety of immune cells, while eicosanoids are key mediators of allergic inflammation. These findings support the possibility that these metabolites are not only biomarkers of remission but may reflect pathophysiologic mechanisms of remission. Our work has been supported by other studies finding differences in the stool metabolome of children on OIT, including amino acids, fatty acids, and bile acids.^24, 25^

Given the evidence that remission is highly prevalent in younger children,^9, 11, 15, 16^ our primary objective was to identify the metabolites associated with remission in young children. Because the DEVIL OIT study of infants and toddlers has the highest 1-month remission rates reported,^11^ we used banked plasma samples from *DEVIL* to study metabolomic pathways that changed over time on OIT and were discrepant between OIT outcomes of remission versus non-remission. Additionally, we sought to validate our prior work of metabolomic profiles of OIT based on enriched pathways from previous analyses.^13^

## Methods

We analyzed metabolomics data from the *DEVIL* OIT clinical trial in young children (9-36 months), conducted at University of North Carolina, Chapel Hill & Duke University (**Figure 1A**).^11^ To validate metabolomic pathways altered during OIT and between remission, we compared our findings in the *DEVIL* study to previously generated metabolomics data and analyses from our comparison cohort, the *PNOIT* study, conducted at Massachusetts General Hospital in older (aged 7-13 years) children (**Figure 1B**).^13, 26^ Analysis of clinical data from *DEVIL* and *PNOIT,* and metabolomics data of PNOIT have been previously described.^11, 13, 26^ General information about the cohorts, metabolomics profiling, and regression and other analysis methods are described below, and additional details can be found in the supplemental methods.

**Figure 1.**
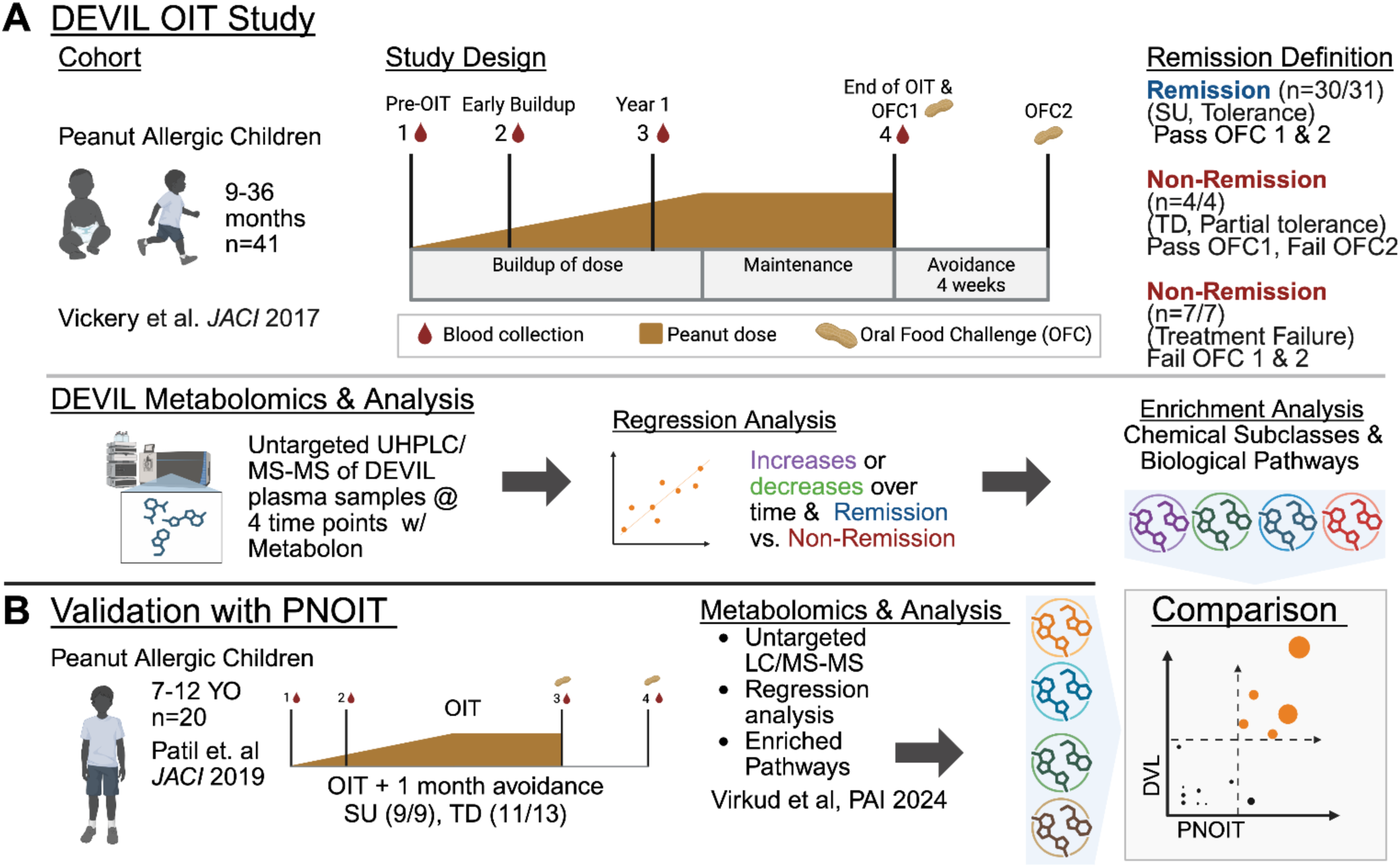
(A) Overview of *DEVIL* cohort study design. To study the metabolomics of peanut OIT we used the *DEVIL* interventional OIT trial.^11^ Participants underwent buildup periods tailored to the patient as this was the first study of children of such young age (1-4YO, n=41). They also underwent maintenance, and 4 weeks of avoidance where it was determined which participants could maintain protection (remission) and consume a full serving of peanut after avoidance and those who lost protection (non-remission). Participants who were unable to pass the oral challenge at end of therapy or withdrew due to adverse events were also classified as non-remission. Untargeted metabolomics was performed on plasma samples from 4 timepoints during OIT to determine which metabolites changed over time on OIT and which metabolites differed by treatment outcomes of remission or non-remission. (B) Validation of *DEVIL* with the *PNOIT* OIT trial. We used a comparison cohort (*PNOIT*) ^13^ to confirm the metabolomic findings from *DEVIL* and narrow down the metabolites most likely predictive of remission or non-remission by comparing enriched chemical subclasses and biological pathways between *PNOIT* and *DEVIL*. Figure 1 created with biorender.com.

### Clinical Cohorts

#### DEVIL

Peanut-allergic children aged 9-36 months (n=41) were enrolled in an interventional peanut OIT trial.^11^ The trial consisted of three main phases, a personalized buildup period of administering gradually increasing daily doses of peanut protein, a maintenance period where patients received a consistent daily dose of peanut protein, and a 1-month avoidance period where no peanut protein was administered. Participants were randomized to two different maintenance doses (300 v 3000 mg). Blood samples were collected in clinic at baseline, early buildup,12 months, and end of therapy.

Double-blind, placebo-controlled oral food challenges (5000 mg of peanut protein) were administered to assess clinical outcomes of remission or non-remission (transient desensitization or treatment failure). Remission is defined as passing both the end of therapy and end of avoidance food challenges, non-remission is defined as reacting to any of those two challenges or withdrawing early due to adverse events from the treatment (Figure 1A).

#### PNOIT

Peanut-allergic children (aged 7-12, n=20) were enrolled in an open-label interventional peanut OIT trial to increase patient’s tolerance to peanut allergen (Figure 1B).^13, 26^ Participants underwent a 44 week long buildup, 12 week maintenance period (dose 4000 mg), and 1 month avoidance period. Remission was evaluated based on end of therapy and end of avoidance food challenges (5000 mg of peanut protein).

### Metabolomics Analysis

#### DEVIL

Metabolomics profiling was conducted on plasma samples at baseline, early build-up, 12 months, and end of therapy. Samples were analyzed by Metabolon Inc using untargeted metabolomics on an Ultra-High Performance Liquid Chromatography (UHPLC) tandem Mass Spectrometry system. Metabolites were labeled with chemical names and Human Metabolome Data base identifiers where available. Chemical subclasses specific to Metabolon’s proprietary database were also provided and for the following analyses are referred to as “lab-identified subclasses”. Following Metabolon’s metabolite identification, we utilized our QC pipeline to further clean the metabolite data. Samples with high levels of missing data were removed, and KNN imputation was used to impute missing metabolite concentrations based off the levels of similar metabolites.^27^ Following imputation of missing metabolites, all features were log-transformed for normalization and pareto-scaled to reduce variation in metabolomic features.

#### PNOIT

Plasma samples (at baseline, early buildup and end of OIT) from participants who achieved transient desensitization and sustained unresponsiveness (n=20) were analyzed by the Broad Institute using untargeted metabolomics and a Liquid Chromatography tandem Mass Spectrometry system. Five quality control (QC) samples were run for reproducibility, and metabolites with an elevated coefficient of variation based on the QC samples were removed. As described above, metabolites with elevated missingness were removed, and missing values were imputed (half of the minimum value detected), and features were log-transformed and pareto-scaled.

### Statistical Analysis

Metabolomic profiles from plasma samples collected through end of therapy, were included in our regression modeling. Given the differences in time within the DEVIL study and compared to the PNOIT study, time was defined by stage (baseline, early buildup, 12 months, and end of OIT). All analyses were performed in R version 4.4.2.^28^

#### Regression analyses

Two sets of regression analyses were performed: (1) models to evaluate metabolites changing over time (time-series models) and (2) models to evaluate metabolites that differed between remission and non-remission. The metabolites that changed significantly over time were identified using generalized linear models (GLM) for each metabolite and were adjusted for age at enrollment, and treatment group. Coefficients from these time-series models, represent the change in pareto-scaled metabolite concentration per month on OIT. For remission outcomes, the odds of developing remission based on metabolite level was estimated using a series of logistic regression models for each metabolite, adjusted for age, treatment group, and timepoint. Odds ratios calculated from logistic regression model coefficients represent the likelihood of developing remission based on metabolite levels, adjusted for time and age at enrollment. All models were adjusted for the treatment group, to account for any differences due to the final maintenance dose.

#### Chemical subclass & Pathway enrichment analysis

We used the Relational database of Metabolomic Pathways (RAMP) to identify “chemical subclasses” based on the HMDB ClassyFire classification system. Using RaMP, we performed two types of enrichment analyses: chemical subclasses enrichment, and biological pathway enrichment based on a compiled library of biological pathways.

Given the hypothesis generating nature of this work, metabolites with a nominal p-value <0.05 in the regression models, were selected for enrichment analysis. False discovery rates (q<0.05) were used for multiplicity correction for all enrichment analysis. Enrichment scores of chemical subclasses and biological pathways were calculated based on the number of observed vs. the number of expected metabolites, using RaMP v3.0.2 (RaMP-DB R package) to identify significantly overrepresented metabolite subclasses and biological pathways.^29, 30^

#### Comparison of PNOIT & DEVIL

Metabolite chemical subclasses and biological pathways were compared between the two cohorts and considered replicated if the FDR from the RaMP based enrichment analysis was significant (q<0.05) in both DEVIL and PNOIT (Figure 1B). Our primary analyses here focused on the subclasses of metabolites that replicated in both *DEVIL* and *PNOIT*, but all associated metabolites and pathways can be found in the supplemental figures.

## Results

### Demographic Summary of the *DEVIL* OIT Cohort

We assessed metabolomics data from 41 children. A majority of the children were male (68.3%), all were white (100%), and ranged in age from 17-41 months at start of OIT **(Table S1)**. A majority had other atopic conditions, most prominently atopic dermatitis over asthma and allergic rhinitis, consistent with the young age of the population. Thirty (73.2%) of the children achieved remission. Of the remaining children who were classified as non-remission, 4 (9.8%) achieved transient desensitization, and 4 (9.8%) failed the desensitization challenge and were considered treatment failures, and 3 (7.3%) withdrew before the end of therapy due to adverse events. The only difference in the remission versus non-remission groups was the lower IgE in the participants who went on to develop remission; there were no demographic differences otherwise. Compared to the *PNOIT* cohort profiled in prior analyses,^13^ the *DEVIL* population was younger (*PNOIT* age range 7-12yo), more female (31.7%; *PNOIT*=25%), more white (100%; *PNOIT*=80%), and a larger proportion of children achieved remission (0.73; *PNOIT*=0.45).

### Metabolomic Trends Over Time on OIT

Of the 881 metabolites, 194 (22%) were associated with time on OIT among all participants, after adjusting for age at enrollment and treatment group (**Figure S1**). These metabolites primarily included lipids and amino acids, comprising 42 lab-identified subclasses (Figure 2A). Of the 194 metabolites significantly associated with time on OIT, 166 (85.6%) were identified by RaMP for enrichment analysis. There were 4 overlapping subclasses that were associated with time on OIT between the *DEVIL* and *PNOIT* cohorts, and 83 biological pathways that were also significantly enriched in both cohorts (Figure 2B**, Figure S2, & Tables S2 & S3**). Based on all of these methods, the metabolite classes most significantly associated with changes over time on OIT included glycerophospholipids and amino acids.

**Figure 2.**
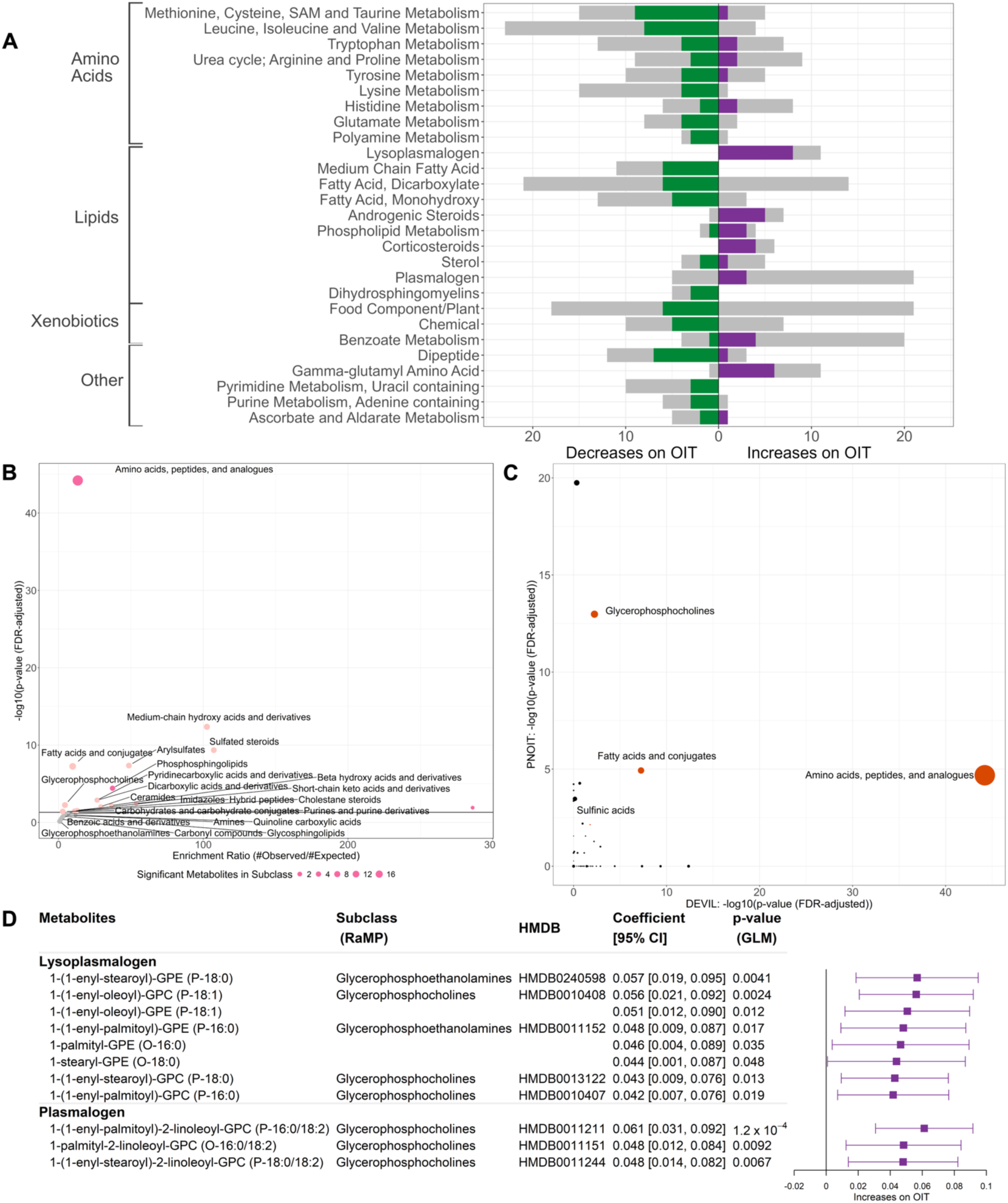
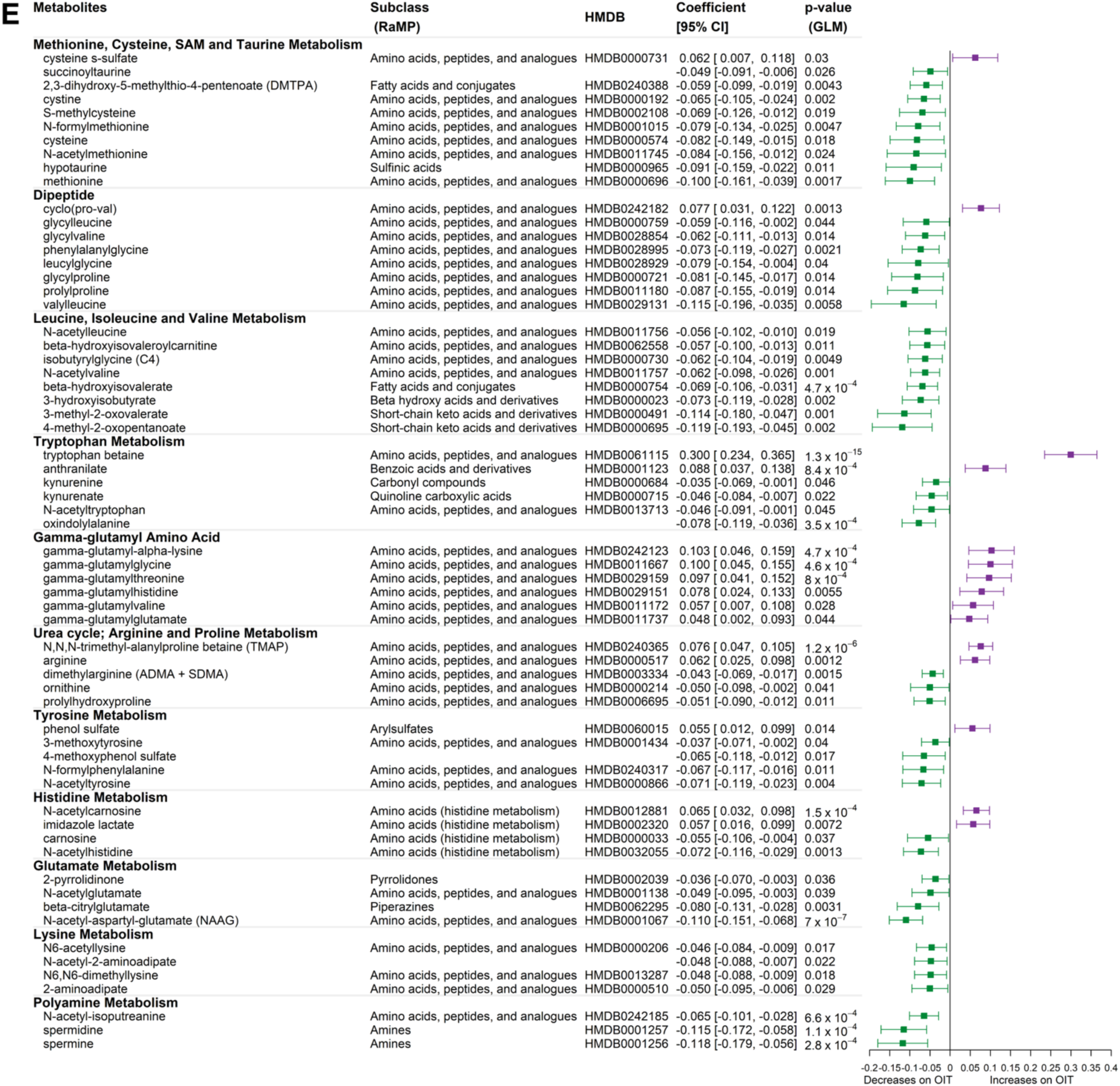
Metabolite changes over time on OIT. Metabolites that increase over time on OIT are in purple and decrease over time on OIT are in green. **A)** Lab-identified chemical subclasses of significantly associated metabolites (included if at least 3+ significant metabolites within the subclass). Grey bars depict the number of detected metabolites in a given class, including non-significant associations, to represent potential annotation biases. **B**) Enrichment of metabolites by RAMP-based chemical subclass. The enrichment ratio (#observed metabolites/#expected based on RaMP database) of metabolite subclasses plotted by the FDR-adjusted enrichment p-value. Dark pink represents subclasses or pathways linked to food allergy in prior publications. **C)** The chemical subclasses significantly enriched in both *DEVIL* and *PNOIT* are indicated in orange. Size of dot reflects the enrichment ratio**. (D & E**) Individual glycerophospholipid (**D**) & amino acid (**E**) metabolites and their association with time on OIT, adjusted for age at enrollment. The coefficient represents the scaled change of metabolite concentration over time on OIT. In Figure 2A, 2D and 2E, we have retained only metabolite subclasses with greater than 3 significant metabolites. All metabolites significantly associated with time on OIT or remission status can be found in **Figure S1**.

### Glycerophospholipids over time on OIT

Glycerophospholipids comprise a group of phospholipid metabolites, which can be classified based on chemical structure of the linkage of their glycerol backbone (e.g. plasmalogens, lysoplasmalogens) or their head-group (e.g. glycerophosphocholines & glycerophosphoethanolamines). Among the lipids, the most abundant lab-identified chemical subclasses associated with time were lysoplasmalogens (Figure 2A). Enrichment analysis showed enrichment of the chemical subclass glycerophosphocholines (q=0.0058; ER=4.4) (Figure 2B), which was one of only 4 chemical subclasses replicated in the PNOIT cohort as well (q=1.05×10^-13^) (Figure 2C). All glycerophospholipids increased over time on OIT [coefficient range=0.042 to 0.061;p-range=1.2×10^-4^-0.048] (Figure 2D).

Closely related to glycerophospholipids, there was also enrichment of sphingolipids including the chemical subclasses of phosphosphingolipids (q=3.8×10^-5^; ER=37.3) (**Table S4**), and the biological pathways sphingolipid metabolism (q=0.000293), and glycosphingolipid catabolism (q=0.012) (**Figure S3 & Table S5**).

### Amino Acids changing over time on OIT

Amino acids were highly abundant among metabolites associated with time, including the lab-identified subclasses of Methionine, Cysteine, SAM, and Taurine Metabolism; Leucine, Isoleucine, and Valine Metabolism; Tryptophan Metabolism; and Urea cycle, Arginine, and proline Metabolism (Figure 2A).

Using RaMP, Amino acids, peptides and analogues (q=6.1×10^-45^, enrichment ratio (ER)=13.3;) was the most significantly enriched chemical subclass, and it was also the top subclass replicated in the PNOIT cohort. (q=2.06×10^-5^) (Figure 2B **& 2C**). Among the biological pathways enriched in both DEVIL & PNOIT, the top pathways included Urea cycle disorders (*DEVIL* q-range=2.55×10^-8^; *PNOIT* q-range=2.06×10^-9^), TCA cycle (*DEVIL* q =0.020; *PNOIT* q=1.29×10^-5^), and Amino acid metabolism pathways (*DEVIL* q =0.00014; *PNOIT* q-range=2.06×10^-9^) (**Figure S2, Table S2 & S3**). Most amino acids decreased over time on OIT [coefficient range= −0.118 to −0.035; p-range=7×10^-7^-0.045], but some metabolites belonging to subclasses such as tryptophan and gamma-glutamyl amino acids increased over time on OIT [coefficient range=0.048-0.30; p-range1.3×10^-15^-0.044 (Figure 2E).

### Differences in the Metabolome between OIT Remission Status

Out of 881 identifiable metabolites, 109 (12.4%) were significantly different between participants who achieved remission and those who did not, after adjusting for time on OIT and age at enrollment. These metabolites comprised 42 subclasses **(Figure S4, Table S6).** Of the 109 metabolites significantly associated with remission status, 84 (77.1%) could be matched to RaMP. In association with remission status, there were 5 overlapping enriched subclasses and 22 biological pathways between the *DEVIL* (**Figure S5, Table S7)** and *PNOIT* cohorts, (Figure 3C**, Tables S8 & S9, Figure S6**). Based on all of these methods, the subclasses/pathways most significantly associated with remission status included glycerophospholipids, amino acids, and bile acids.

**Figure 3.**
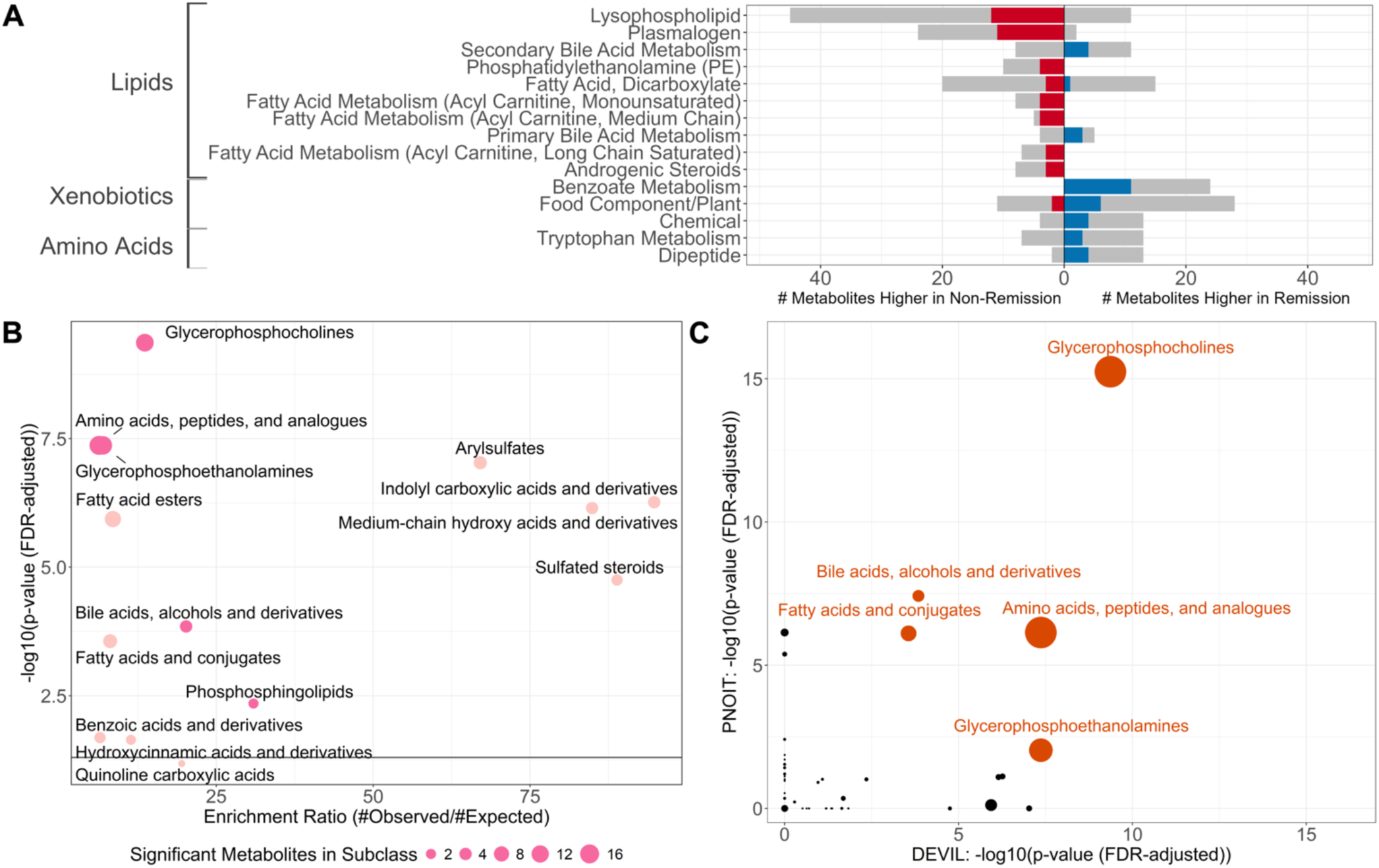
Metabolite differences by OIT remission status. **A)** Lab-identified subclasses associated with remission (blue) and non-remission (red). Grey bars represent the number of detected metabolites in a specific subclass, including non-significant associations, to depict potential annotation bias. **B)** Enrichment of metabolites by RAMP-based chemical subclass. The enrichment ratio (#observed metabolites/#expected based on RaMP database) of metabolite subclasses plotted by the FDR-adjusted enrichment p-value. Dark pink represents subclasses or pathways linked to food allergy in prior publications. **C)** The chemical subclasses significantly enriched in both *DEVIL* and *PNOIT* among metabolites significantly associated with remission outcome are indicated in orange. Size of dot indicates enrichment ratio. In Figure 3A, we have retained only metabolite subclasses with greater than 3 significant metabolites, all metabolites significantly associated with time on OIT or remission status can be found in **Figure S4**.

<u>Remission and Phospholipids (Glycerophospholipids, Plasmalogens, & Sphingolipids)</u> Glycerophospholipids, including lysophospholipids, plasmalogens, and phosphatidylethanolamines, were among the lab-identified metabolite classes most associated with remission versus non-remission (Figure 3A). Based on RAMP chemical subclasses enrichment analysis, glycerophosphocholines (q=4.30×10^-10^, enrichment ratio=13.6) and glycerophosphoethanolamines (q=4.28×10^-8^, enrichment ratio=6.9) were among the top enriched subclasses in both DEVIL and in PNOIT (Figure 3B **& 3C**). All glycerophospholipid metabolites significantly associated with remission status were higher in non-remission (range of OR:0.04-0.26, range of p=5.9e-4-0.048), and a majority of non-significant associations trended towards non-remission as well (Figure 3A **& 4A & 4B**). While some metabolites within these classes were similar at baseline between remission and non-remission, overall we observed the same trends that glycerophospholipid concentrations remained relatively stable in the remission group and increased over time in the non-remission group (Figure 4B).

**Figure 4.**
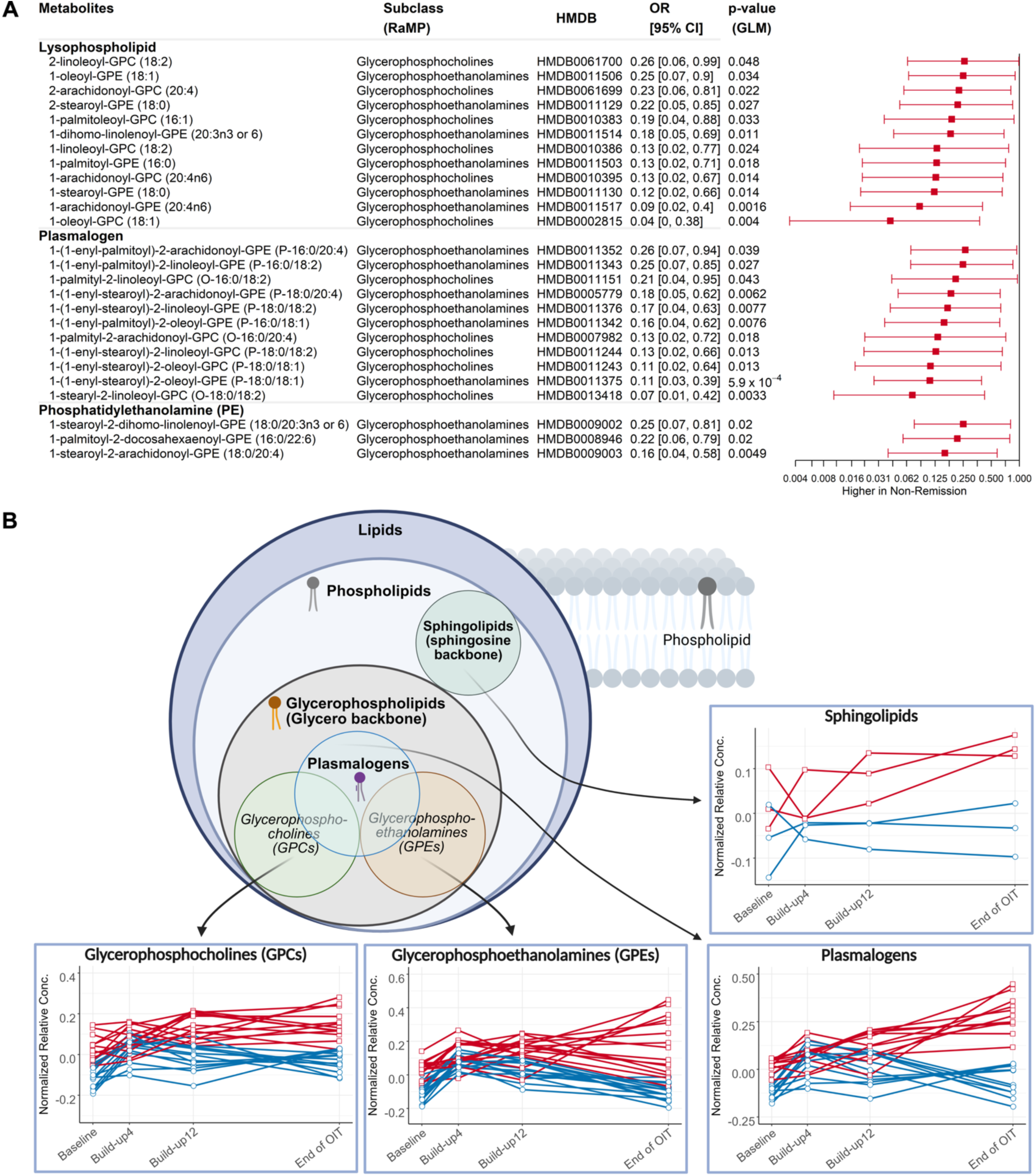
Glycerophospholipids and Remission. **A)** Individual glycerophospholipid metabolites and their association with OIT remission status, adjusted for time, age at enrollment, and dose. The odds ratio (OR) represents the likelihood of developing remission (>1) or non-remission (<1) on OIT for each scaled unit metabolite concentration. **B)** Overview of glycerophospholipid groupings and individual metabolite trajectories by remission status during OIT. Glycerophospholipid metabolites associated with remission on OIT are depicted in blue, while metabolites associated with non-remission are depicted in red. Plasmalogens are a functional group consisting of a subset of GPCs and GPEs, the plasmalogens figure consists of the GPCs and GPEs which are also part of the plasmalogen functional group. In Figure 4A, we have retained only metabolite subclasses with greater than 3 significant metabolites, all metabolites significantly associated with time on OIT or remission status can be found in **Figure S4**. Figure 4B created with biorender.com

### Remission and Amino Acids

A large number of amino acids were associated with remission status (21 out of 108 associated metabolites) spanning 12 different lab-identified subclasses, though only the tryptophan metabolism and dipeptide subclasses had at least 3 metabolites within the group (Figure 3A**, Figure S4**). Based on RAMP pathway analysis, amino acids were among the top highly significant subclasses in both DEVIL (q=4.28×10^-8^) and PNOIT (q=7.2×10^-7^) (Figure 3B **& 3C, Table S8**). Top overlapping biological pathways in DEVIL and replicated in PNOIT included urea cycle disorders (DEVIL q=0.0004; PNOIT q=2.32×10^-9^), creatine metabolism (DEVIL q=0.0007, PNOIT q=9.76×10^-6^), and amino acid pathways (DEVIL q=0.0017; PNOIT q=3.01×10^-12^) (**Figure S6, Table S9**). Of the two subgroups with sufficient replication (3+ metabolites), all of the tryptophan metabolism and dipeptide metabolites were higher in the remission group compared to the non-remission group (Figure 5).

**Figure 5.**
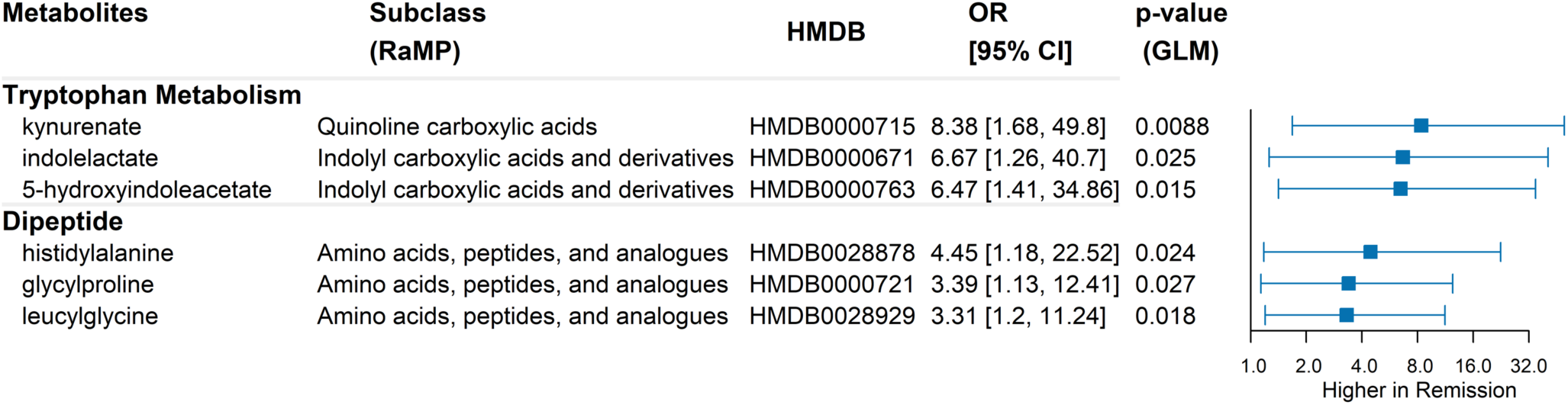
Individual amino acid metabolites and their association with remission status, adjusted for time, age at enrollment, and dose. The odds ratio (OR) represents the likelihood of developing remission (>1) or non-remission (<1) on OIT for each scaled unit metabolite concentration. Only metabolite subclasses with greater than 3 significant metabolites are included. All metabolites significantly associated with time on OIT or remission status can be found in **Figure S4**.

### Remission and Bile Acids

Bile acids, including both primary and secondary bile acids were among the most abundantly reported lab-identified subclasses (Figure 3A). RaMP based enrichment analyses confirmed that the bile acids, alcohols and derivatives subclass (q=0.00014, enrichment ratio=20.2) was highly enriched in DEVIL and also overlapped with PNOIT (q=3.8×10^-8^) (Figure 3B **& 3C**). In DEVIL, all of the bile acids were higher in the remission group compared to non-remission (Figure 6A). Individual trajectories varied by metabolite with the cheno-deoxycholate derived bile acids having similar levels prior to OIT and diverging after OIT (muricholates, taurolithocholate 3-sulfate, and glycochenodeoxycholate glucoronide), while the cholic acid derived bile acids differed even at baseline (taurodeoxycholate, taurocholenate, and glycodeoxycholate) (Figure 6B).

**Figure 6.**
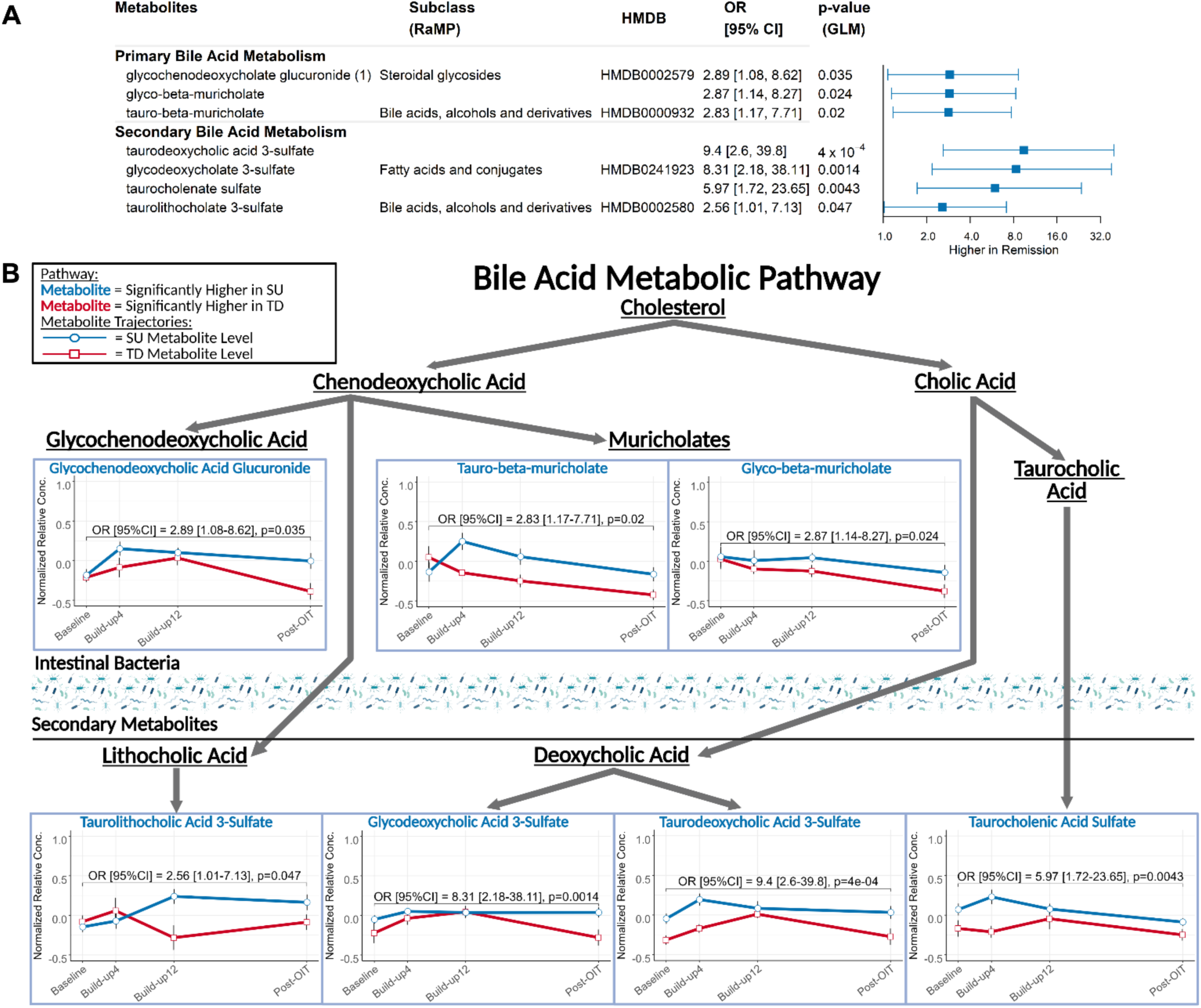
Bile Acids and Remission. **A)** Individual bile acid metabolites and their association with remission, adjusted for time, age at enrollment, and dose. The odds ratio (OR) represents the likelihood of developing remission (>1) or non-remission (<1) on OIT for each scaled unit metabolite concentration. Only metabolite subclasses with greater than 3 significant metabolites are included here. All metabolites significantly associated with time on OIT or remission status can be found in **Figure S4**. **B)** Overview of bile acid pathways and glycerophospholipid groupings and individual metabolite trajectories by remission status during OIT. Bile acid pathway metabolites associated with remission on OIT are depicted in blue, while metabolites associated with non-remission are depicted in red. Figure 6B created with biorender.com.

## Discussion

Using samples from the *DEVIL* clinical OIT trial, we aimed to identify metabolites associated with remission in young children and validate these findings with our prior work in the *PNOIT* OIT trial in older children. To our knowledge, this is the first study of plasma metabolomics in younger children and comparison of the metabolomics of younger and older children on OIT. We found that in young children on peanut OIT, bile acids and amino acids were higher in remission, compared to glycerophospholipids which were higher in the non-remission group. Over time on OIT, amino acids were generally decreasing and glycerophospholipids were increasing over time on OIT.

### Bile Acids

We found that bile acids were were all higher in remission compared to non-remission. Early literature shows differences in bile acids when comparing food allergic children to non-food allergic controls.^13, 20, 21, 31^ In our prior work on *PNOIT*, we reported that most bile acids were higher in non-remission, while lithocholate derivatives were higher in remission group.^13^ Here, we also found the taurolithocholate derivative was elevated in remission, but other bile acids were also elevated in remission. These differences may be due to the younger age of the cohort or may be related to differences in the individual metabolites detected between the two cohorts.

Because of the role of gut microbiota in modifying bile acids, the plasma metabolome is significantly affected by the intestinal microbiome. In a preprint of a study on the fecal microbiome and metabolome of young children on OIT, children with remission had distinct microbial patterns related to bile acid deconjugation.^24^ This study also demonstrated higher levels of certain lithocholate derivatives, including taurolithocholates, in remission versus non-remission.^24^

Prior research demonstrates bile acids have immunomodulatory roles on a variety of immune cells, including T cells, dendritic cells, B cells, and more. ^32, 33^ Certain lithocholate derivatives have been shown to block Th17 differentiation and enhance T-regulatory cell differentiation and function.^34–37^ Given the role of these T cell subsets in remission during OIT, this research provides a potential mechanism for the development of remission while on OIT.^38–40^

Combined, these studies imply that bile acids, and especially lithocholates, could not only be biomarkers of remission but could play a role in the mechanism of remission, connecting the stool microbiome to stool and plasma metabolite profiles that thereby influence immune cell phenotypes. These experimental findings need to be validated in a larger cohort, but it is significant that we see the replication of bile acid enrichment between independent pediatric food allergy cohorts of different ages.

### Glycerophospholipids (Plasmalogens, Glycerophosphocholines & Glycerophosphoethanolamines) and Sphingolipids)

In *DEVIL*, glycerophospholipids, including plasmalogens, increased over time on OIT, though this appeared to be largely driven by increases in the non-remission group and stable levels in the remission group (Figure 4). While both the *PNOIT* and *DEVIL* studies showed enrichment of glycerophosphocholines and glycerophosphoethanolamines, in *PNOIT* the glycerophospholipids are more discrepant, with only 60% of the metabolites higher in the non-remission group, which may be due to either differences in the two cohorts or the individual metabolites detected.^13^

Glycerophospholipids are a class of sphingolipids, that include plasmalogens, a chemical subclass that has been associated with atopic diseases.^41–43^ Plasmalogens are comprised of glycerophosphocholines and glycerophosphoethanolamines that contain a vinyl-ether bond at the sn-1 position of the glycerol backbone.^44^ Plasmalogens comprise 30-40% of the glycerophospholipids identified as higher in non-remission in both the *DEVIL* and *PNOIT* studies. A study of food allergy prevalence and metabolomics found increased levels of plasmalogens in the serum of children with multiple food allergies.^20^ These studies support further study of the role of plasmalogens and other glycerophospholipids in food allergy and remission on OIT.

### Amino Acids (Urea Cycle & Tryptophan) Metabolites

In both the *DEVIL* and *PNOIT* studies, amino acid, tryptophan and urea cycle metabolite pathways were observed as significantly enriched among metabolites significantly associated with time on OIT and with remission. We and others have shown associations of amino acid metabolites, particularly histidine metabolites, with food allergy. More recently, altered fecal amino acid metabolism was described between remission and non-remission groups during OIT, which may have implications of plasma amino acid profiles.^24^

### Limitations

This study is limited by small sample sizes, and the differences between the 2 cohorts, including both study design and metabolomics profiling. While the overall structure, avoidance periods, and food challenge doses were the same, the differences in maintenance doses and build-up periods could have affected the findings. Metabolomic profiling was conducted by two different metabolomics analysis groups (Metabolon Inc and the Broad Institute). Individual differences between laboratories is known to cause discrepancies in the uniformity of metabolite detection, but not the detection of individual metabolites. The lack of racial diversity in the individual studies does affect the generalizability of these results. Because we do not have dietary data, we are unable to assess the influence of a variable infant & toddler diet on these metabolomic findings. In general, most of these limitations bias our findings towards the null. Furthermore, the replication of important findings between these studies despite their differences and small sample sizes bolsters our confidence in the findings. It will be important to replicate these findings in larger and more diverse cohorts that reflect the demographics of the food-allergic population.

## Conclusion

This study provides a comparison of metabolites associated with time on OIT and remission status between two pediatric OIT cohorts of different ages. We found higher levels of bile acids, including lithocholate derivatives, among the children who developed remission, and higher levels of glycerophospholipids among the children who developed non-remission, increasing over time on therapy. Many of these metabolites, including bile acids and amino acids are known to have immunomodulatory roles on key immune subsets that have been shown to be involved in remission on OIT, suggesting possible mechanisms of remission. With further study, glycerophospholipids, amino acids, and bile acids have the potential to be used as biomarkers in determining the likelihood of developing tolerance while on OIT or have the potential to identify new mechanisms important in the modification of tolerance in food allergy.

## Data Availability

All data produced in the present study will be made available upon publication of this work.

