## Supplemental Methods for "Association of bile acids, amino acids, and glycerophospholipid metabolites with food-allergic outcomes in children on peanut oral immunotherapy"

We analyzed metabolomics data from the Determining the Efficacy and Value of Immunotherapy on the Likelihood of Peanut Tolerance (DEVIL) OIT clinical trials in young children (9-36 months old), conducted at University of North Carolina, Chapel Hill & Duke University. [1] To validate metabolomic pathways altered during OIT and between remission, we compared our findings in the DEVIL study to previously generated metabolomics data and analyses from our comparison cohort, the *PNOIT* study, conducted at Massachusetts General Hospital in older (7-12 yo) children.[2, 3] Analysis of clinical data from *DEVIL* and *PNOIT* have been previously described.[1-3] For both studies, IRB approval from host institutions was received. Additionally, parental consent on behalf of all children and assent, where age appropriate, were obtained for all participants in both studies.

<u>Cohort Descriptions</u>

*DEVIL:* Children aged 9-36 months (n=41) were enrolled in an interventional peanut OIT trial (ClinicalTrials.gov ID: NCT00932828). Children were randomized to one of two arms based on the final maintenance dose, either 300 or 3000 mg. The trial consisted of three main phases, a buildup period of administering gradually increasing daily doses of peanut protein, a maintenance period where patients received a consistent daily dose of peanut protein, and a 1-month avoidance period where no peanut protein was administered. Blood samples were collected in clinic at baseline (prior to starting therapy), early buildup (~4 months, an immunologically active period on OIT), maintenance (12 months), end of therapy (end of maintenance), and end of avoidance (1 month after end of therapy). Double-blind, placebo-controlled oral food challenges (5 grams of peanut protein) were administered at end of therapy and end of avoidance. As this was the first OIT study in this young age of children, the length of buildup was tailored to each child and treatment lengths ranged from 21 to 65 months. Study outcomes included transient desensitization, defined as passing the end of therapy oral food challenge, and 1 month remission (also known as sustained unresponsiveness), defined as passing both the end of therapy and end of avoidance oral food challenge.

*PNOIT:* Children (aged 7-12, n=20) were enrolled in an open-label interventional peanut OIT trial to increase patient's tolerance to peanut allergen (ClinicalTrials.gov ID: NCT01324401). The study protocol involved giving increasing doses of daily peanut allergen (buildup), followed by a maintenance period of consuming a daily dose of 4g, followed by a final 1-month period of peanut allergen avoidance. Oral food challenges (5 g of peanut protein) were performed at the end of maintenance (end of therapy) and at the end of avoidance to determine remission status. The food challenge at the end of avoidance was a double-blind placebo-controlled food challenge. Blood samples were collected from participants at baseline, early build-up (when adverse events were occurring at the highest rate), at the end of maintenance (end of therapy), and following the 1-month period of avoidance (end of avoidance). Blood samples were collected during clinic visits and prior to conducting oral food challenges where applicable. Additional details regarding study design are available in the original publications of the *PNOIT* trial.[3] Food allergy cases were identified and eligible for study participation based on observation of objective symptoms after peanut ingestion, and either a peanut-specific IgE >10 kU/L or elevated peanut-specific skin prick test >8 mm. Food allergy presence was further confirmed when all patients demonstrated symptoms of IgE-mediated food allergy during buildup.

<u>Metabolomics Analysis</u>

*DEVIL*: Plasma samples were stored at -20C in the UNC FAI laboratory freezers following collection until being transferred to Metabolon. Metabolomics profiling was conducted on plasma samples with sufficient volume (>100 microliters) for the baseline, early build-up, 12 months, end of therapy, and end of avoidance timepoints. Samples were analyzed by Metabolon Inc using untargeted metabolomics on an Ultra-High Performance Liquid Chromatography (UHPLC) tandem Mass Spec system. Metabolon identified individual metabolites based on mass-to-charge ratio, LC retention time, and any other features captured by Metabolon's proprietary and automated mass-spec data extraction and reduction software. Metabolon's proprietary reference library of metabolites was used to identify metabolite subclasses (referred to in the manuscript as 'lab-identified subclasses', using their automated proprietary software and verified by data curators. Following metabolite identification, we utilized our QC pipeline to further clean the metabolite data. Samples with <40% missing estimated metabolite concentrations were included in further analysis. KNN imputation was used to impute missing metabolite concentrations based off the levels of similar metabolites.[4]

*PNOIT*: Plasma samples from participants who achieved transient desensitization and sustained unresponsiveness (n=20) were analyzed by the Broad Institute using untargeted metabolomics and tandem Liquid Chromatography Mass Spec. Five quality control (QC) samples were run for reproducibility, and metabolites with a coefficient of variation > 25% were removed. Metabolites with missing data across >10% of the samples were removed, and then remaining missing values were imputed using half of the minimum value detected for each given metabolite.

Following imputation of missing metabolites, all features were log-transformed for normalization and *pareto*-scaled to reduce the variation in fold change differences between the features. Due to the nature of untargeted metabolomics, where we are dealing with relative metabolite concentrations, it is beneficial to adjust for the means and variances (*pareto*-scaling) of individual metabolites. Unnamed (unidentified) metabolites were removed from our analyses.

Statistical Analysis

*DEVIL*: Metabolomic profiles from plasma samples collected through end of therapy, were included in our regression modeling. End of avoidance profiles were excluded to focus on metabolomic trends while on therapy. All analyses were performed using R version 4.4.2.[5]

The metabolites that changed significantly over time were identified using GLM models for each metabolite and were adjusted for age. Coefficients from these models (figure 2), represent the change in pareto-scaled metabolite per month on OIT, adjusted for age at enrollment, and treatment group.

$$\mathbf{y} = \boldsymbol{\beta}_0 + \boldsymbol{\beta}_1 * \boldsymbol{time} + \boldsymbol{\beta}_2 * \boldsymbol{age} + \boldsymbol{\beta}_3 * \boldsymbol{treatment\ group} + \boldsymbol{\varepsilon}$$

$\mathbf{y}$ = individual metabolite concentrations
$\boldsymbol{time}$ = time on OIT (4 timepoints used: pre-OIT (baseline), early build-up, 12 months, and post-OIT)
$\boldsymbol{age}$ = age at the time of enrollment in months
$\boldsymbol{treatment\ group}$ = treatment group, based on randomized maintenance dose

Additionally, the odds of developing remission based on metabolite level was estimated using a series of logistic regression models for each metabolite, adjusted for age, timepoint, and treatment group. Odds ratios calculated from logistic regression model coefficients represent the likelihood of developing remission based on metabolite levels, adjusted for time and age at enrollment. An odds ratio greater than one represents an increased likelihood of developing remission, while an odds ratio less than one estimates an increased likelihood of not developing remission.

$$\log\left(\frac{P}{1-P}\right) = \beta_0 + \beta_1 * metabolite_i + \beta_2 * time + \beta_3 * age + \beta_4 * treatment\ group + \varepsilon$$

$\mathbf{P}$ = probability of having remission versus non-remission
$metabolite_i$ = individual metabolite concentrations
$time$ = time on OIT (4 timepoints used: pre-OIT (baseline), early build-up, 12 months, and post-OIT)
$age$ = age at the time of enrollment in months
$treatment\ group$ = treatment group, based on randomized maintenance dose

More complex modeling, including mixed effects models (participant as random effects, and time and/or outcome as fixed effects) and models with interaction terms for time, were evaluated but could not be performed due to lack of model convergence with the given sample sizes.

Human Metabolome Data Base (HMDB) identifiers of metabolites significantly associated with time on OIT or remission were analyzed by enrichment analysis using RaMP to identify significantly overrepresented metabolite subclasses and biological pathways. Metabolite subclass identification based on HMDB numbers, biological pathway enrichment, and chemical subclass enrichment analyses were performed using RaMPv3.0.2 (RaMP-DB R package). [6, 7] Individual metabolites were considered significant based on α=0.05 to capture all potentially relevant subclasses and biological pathways, while enrichment analyses results were reported using a Benjamini-Hochberg corrected significance of q<0.05. We identified several subclasses of metabolites that have demonstrated consistent association with the PNOIT cohort (based on our comparison analysis, described below) and we have highlighted these subclasses in our analyses.

*Comparison of PNOIT & DEVIL*: Metabolite chemical subclasses and biological pathways that were enriched in the PNOIT enrichment analysis were selected.[2] Subclasses and pathways were defined as replicated, based on subclass name and pathway ID respectively, if they were significant (q<0.05) in both cohorts.
