## Supplemental Figures and Tables for "Association of bile acids, amino acids, and glycerophospholipid metabolites with food-allergic outcomes in children on peanut oral immunotherapy"

**Supplemental Figures & Tables**

Supplemental Table 1. Demographic summary of DEVIL peanut OIT trial participants. Only participants whose plasma samples were able to be analyzed for metabolomics were included here.

| | *level* | **Overall (%)** | **Non-Remission (%)** | **Remission (%)** | **p** |
|---|---|---|---|---|---|
| *n* | | *41* | *11* | *30* | |
| **Sex** | *Female* | 13 ( 31.7) | 4 ( 36.4) | 9 ( 30.0) | 0.99 |
| | *Male* | 28 ( 68.3) | 7 ( 63.6) | 21 ( 70.0) | |
| **Race** | *White* | 41 (100.0) | 11 (100.0) | 30 (100.0) | NA |
| **Median Age in Years [IQR]** | | 2.6 [2.2, 3.0] | 2.9 [2.7, 3.0] | 2.6 [1.9, 3.0] | 0.10 |
| **Median Age in Months [IQR]** | | 32.0 [26.0, 36.0] | 35.0 [32.0, 36.0] | 31.0 [22.8, 35.0] | 0.13 |
| **Asthma** | | 31 ( 75.6) | 10 ( 90.9) | 21 ( 70.0) | 0.33 |
| | *Yes* | 10 ( 24.4) | 1 ( 9.1) | 9 ( 30.0) | |
| **Atopic Dermatitis** | | 12 ( 29.3) | 2 ( 18.2) | 10 ( 33.3) | 0.58 |
| | *Yes* | 29 ( 70.7) | 9 ( 81.8) | 20 ( 66.7) | |
| **Allergic Rhinitis** | | 30 ( 73.2) | 9 ( 81.8) | 21 ( 70.0) | 0.72 |
| | *Yes* | 11 ( 26.8) | 2 ( 18.2) | 9 ( 30.0) | |
| **Median Baseline SPT [IQR]** | | 12.0 [8.0, 16.5] | 14.0 [8.3, 19.3] | 10.5 [8.0, 16.3] | 0.45 |
| **Median Peanut IgE [IQR]** | | 14.1 [4.9, 28.2] | 100.0 [32.8, 100.0] | 13.2 [3.5, 18.9] | 0.042 |

Supplemental Figure 1. Metabolites significantly associated with time on OIT, adjusted for age at enrollment. The coefficient represents the scaled change of metabolite concentration over time on OIT.

| Metabolites | Subclass (RaMP) | HMDB | Coefficient [95% CI] | p-value (GLM) |
|---|---|---|---|---|
| **Methionine, Cysteine, SAM and Taurine Metabolism** | | | | |
| cysteine s-sulfate | Amino acids, peptides, and analogues | HMDB0000731 | 0.062 [0.0066, 0.12] | 0.03 |
| succinoyltaurine | | | -0.049 [-0.091, -0.0064] | 0.026 |
| 2,3-dihydroxy-5-methylthio-4-pentenoate (DMTPA) | Fatty acids and conjugates | HMDB0240388 | -0.059 [-0.099, -0.019] | 0.0043 |
| cystine | Amino acids, peptides, and analogues | HMDB0000192 | -0.065 [-0.11, -0.024] | 0.002 |
| S-methylcysteine | Amino acids, peptides, and analogues | HMDB0002108 | -0.069 [-0.13, -0.012] | 0.019 |
| N-formylmethionine | Amino acids, peptides, and analogues | HMDB0001015 | -0.079 [-0.13, -0.025] | 0.0047 |
| cysteine | Amino acids, peptides, and analogues | HMDB0000574 | -0.082 [-0.15, -0.015] | 0.018 |
| N-acetylmethionine | Amino acids, peptides, and analogues | HMDB0011745 | -0.084 [-0.16, -0.012] | 0.024 |
| hypotaurine | Sulfinic acids | HMDB0000965 | -0.091 [-0.16, -0.022] | 0.011 |
| methionine | Amino acids, peptides, and analogues | HMDB0000696 | -0.1 [-0.16, -0.039] | 0.0017 |
| **Dipeptide** | | | | |
| cyclo(pro-val) | Amino acids, peptides, and analogues | HMDB0242182 | 0.077 [0.031, 0.12] | 0.0013 |
| glycylleucine | Amino acids, peptides, and analogues | HMDB0000759 | -0.059 [-0.12, -0.002] | 0.044 |
| glycylvaline | Amino acids, peptides, and analogues | HMDB0028854 | -0.062 [-0.11, -0.013] | 0.014 |
| phenylalanylglycine | Amino acids, peptides, and analogues | HMDB0028995 | -0.073 [-0.12, -0.027] | 0.0021 |
| leucylglycine | Amino acids, peptides, and analogues | HMDB0028929 | -0.079 [-0.15, -0.0043] | 0.04 |
| glycylproline | Amino acids, peptides, and analogues | HMDB0000721 | -0.081 [-0.15, -0.017] | 0.014 |
| prolylproline | Amino acids, peptides, and analogues | HMDB0011180 | -0.087 [-0.16, -0.019] | 0.014 |
| valylleucine | Amino acids, peptides, and analogues | HMDB0029131 | -0.12 [-0.2, -0.035] | 0.0058 |
| **Lysoplasmalogen** | | | | |
| 1-(1-enyl-stearoyl)-GPE (P-18:0) | Glycerophosphoethanolamines | HMDB0240598 | 0.057 [0.019, 0.095] | 0.0041 |
| 1-(1-enyl-oleoyl)-GPC (P-18:1) | Glycerophosphocholines | HMDB0010408 | 0.056 [0.021, 0.092] | 0.0024 |
| 1-(1-enyl-oleoyl)-GPE (P-18:1) | | | 0.051 [0.012, 0.09] | 0.012 |
| 1-(1-enyl-palmitoyl)-GPE (P-16:0) | Glycerophosphoethanolamines | HMDB0011152 | 0.048 [0.0092, 0.087] | 0.017 |
| 1-palmityl-GPE (O-16:0) | | | 0.046 [0.0037, 0.089] | 0.035 |
| 1-stearyl-GPE (O-18:0) | | | 0.044 [0.00086, 0.087] | 0.048 |
| 1-(1-enyl-stearoyl)-GPC (P-18:0) | Glycerophosphocholines | HMDB0013122 | 0.043 [0.0094, 0.076] | 0.013 |
| 1-(1-enyl-palmitoyl)-GPC (P-16:0) | Glycerophosphocholines | HMDB0010407 | 0.042 [0.0072, 0.076] | 0.019 |
| **Leucine, Isoleucine and Valine Metabolism** | | | | |
| N-acetylleucine | Amino acids, peptides, and analogues | HMDB0011756 | -0.056 [-0.1, -0.0099] | 0.019 |
| beta-hydroxyisovaleroylcarnitine | Amino acids, peptides, and analogues | HMDB0062558 | -0.057 [-0.1, -0.013] | 0.011 |
| isobutyrylglycine (C4) | Amino acids, peptides, and analogues | HMDB0000730 | -0.062 [-0.1, -0.019] | 0.0049 |
| N-acetylvaline | Amino acids, peptides, and analogues | HMDB0011757 | -0.062 [-0.098, -0.026] | 0.001 |
| beta-hydroxyisovalerate | Fatty acids and conjugates | HMDB0000754 | -0.069 [-0.11, -0.031] | $4.7 \times 10^{-4}$ |
| 3-hydroxyisobutyrate | Beta hydroxy acids and derivatives | HMDB0000023 | -0.073 [-0.12, -0.028] | 0.002 |
| 3-methyl-2-oxovalerate | Short-chain keto acids and derivatives | HMDB0000491 | -0.11 [-0.18, -0.047] | 0.001 |
| 4-methyl-2-oxopentanoate | Short-chain keto acids and derivatives | HMDB0000695 | -0.12 [-0.19, -0.045] | 0.002 |
| **Tryptophan Metabolism** | | | | |
| tryptophan betaine | Amino acids, peptides, and analogues | HMDB0061115 | 0.3 [0.23, 0.36] | $1.3 \times 10^{-15}$ |
| anthranilate | Benzoic acids and derivatives | HMDB0001123 | 0.088 [0.037, 0.14] | $8.4 \times 10^{-4}$ |
| kynurenine | Carbonyl compounds | HMDB0000684 | -0.035 [-0.069, -0.00085] | 0.046 |
| kynurenate | Quinoline carboxylic acids | HMDB0000715 | -0.046 [-0.084, -0.007] | 0.022 |
| N-acetyltryptophan | Amino acids, peptides, and analogues | HMDB0013713 | -0.046 [-0.091, -0.0015] | 0.045 |
| oxindolylalanine | | | -0.078 [-0.12, -0.036] | $3.5 \times 10^{-4}$ |
| **Gamma-glutamyl Amino Acid** | | | | |
| gamma-glutamyl-alpha-lysine | Amino acids, peptides, and analogues | HMDB0242123 | 0.1 [0.046, 0.16] | $4.7 \times 10^{-4}$ |
| gamma-glutamylglycine | Amino acids, peptides, and analogues | HMDB0011667 | 0.1 [0.045, 0.15] | $4.6 \times 10^{-4}$ |
| gamma-glutamylthreonine | Amino acids, peptides, and analogues | HMDB0029159 | 0.097 [0.041, 0.15] | $8 \times 10^{-4}$ |
| gamma-glutamylhistidine | Amino acids, peptides, and analogues | HMDB0029151 | 0.078 [0.024, 0.13] | 0.0055 |
| gamma-glutamylvaline | Amino acids, peptides, and analogues | HMDB0011172 | 0.057 [0.0065, 0.11] | 0.028 |
| gamma-glutamylglutamate | Amino acids, peptides, and analogues | HMDB0011737 | 0.048 [0.0017, 0.093] | 0.044 |
| **Fatty Acid, Dicarboxylate** | | | | |
| 2-hydroxyglutarate | Short-chain hydroxy acids and derivatives | HMDB0059655 | -0.048 [-0.087, -0.0096] | 0.016 |
| 3-hydroxydodecanedioate | Medium-chain hydroxy acids and derivatives | HMDB0000413 | -0.055 [-0.1, -0.0054] | 0.031 |
| heptenedioate (C7:1-DC) | | | -0.055 [-0.1, -0.008] | 0.023 |
| sebacate (C10-DC) | Fatty acids and conjugates | HMDB0000792 | -0.057 [-0.11, -0.0053] | 0.032 |
| tridecenedioate (C13:1-DC) | | | -0.059 [-0.12, -0.003] | 0.041 |
| 2-hydroxysebacate | Medium-chain hydroxy acids and derivatives | HMDB0000424 | -0.063 [-0.12, -0.0094] | 0.023 |
| **Food Component/Plant** | | | | |
| tartronate (hydroxymalonate) | Dicarboxylic acids and derivatives | HMDB0035227 | -0.05 [-0.085, -0.015] | 0.0059 |
| mannonate | Amino acids, peptides, and analogues | HMDB0253141 | -0.06 [-0.098, -0.023] | 0.002 |
| phytanate | Diterpenoids | HMDB0000801 | -0.067 [-0.13, -0.0062] | 0.032 |
| 2-keto-3-deoxy-gluconate | Medium-chain keto acids and derivatives | HMDB0001353 | -0.1 [-0.17, -0.034] | 0.0036 |
| 2,8-quinolinediol sulfate | Quinolones and derivatives | HMDB0240658 | -0.11 [-0.16, -0.051] | $3 \times 10^{-4}$ |
| 2-piperidinone | Piperidinones | HMDB0011749 | -0.13 [-0.21, -0.048] | 0.0021 |
| **Medium Chain Fatty Acid** | | | | |
| 10-undecenoate (11:1n1) | Fatty acids and conjugates | HMDB0033724 | -0.057 [-0.11, -0.0046] | 0.035 |
| cis-4-decenoate (10:1n6) | Fatty acids and conjugates | HMDB0004980 | -0.062 [-0.11, -0.013] | 0.014 |
| 5-dodecenoate (12:1n7) | Fatty acids and conjugates | HMDB0000529 | -0.065 [-0.13, -0.0027] | 0.043 |
| caprylate (8:0) | Fatty acids and conjugates | HMDB0000482 | -0.073 [-0.14, -0.01] | 0.024 |
| caprate (10:0) | Fatty acids and conjugates | HMDB0000511 | -0.075 [-0.14, -0.0049] | 0.038 |
| caproate (6:0) | Fatty acids and conjugates | HMDB0000535 | -0.084 [-0.14, -0.03] | 0.0028 |
| **Benzoate Metabolism** | | | | |
| 4-vinylphenol sulfate | Arylsulfates | HMDB0062775 | 0.17 [0.1, 0.23] | $1.8 \times 10^{-6}$ |
| catechol sulfate | Arylsulfates | HMDB0059724 | 0.082 [0.03, 0.13] | 0.0024 |
| guaiacol sulfate | Arylsulfates | HMDB0060013 | 0.081 [0.023, 0.14] | 0.0065 |
| 4-methylcatechol sulfate | Arylsulfates | HMDB0240459 | 0.069 [0.011, 0.13] | 0.022 |
| methyl-4-hydroxybenzoate sulfate | Carbonyl compounds | HMDB0041646 | -0.18 [-0.29, -0.084] | $4.6 \times 10^{-4}$ |
| **Androgenic Steroids** | | | | |
| androsterone sulfate | Sulfated steroids | HMDB0002759 | 0.13 [0.057, 0.19] | $4.3 \times 10^{-4}$ |
| dehydroepiandrosterone sulfate (DHEA-S) | Sulfated steroids | HMDB0001032 | 0.12 [0.053, 0.2] | $8.5 \times 10^{-4}$ |
| androstenediol (3beta,17beta) disulfate (1) | Sulfated steroids | HMDB0240313 | 0.11 [0.051, 0.18] | $6 \times 10^{-4}$ |
| epiandrosterone sulfate | Sulfated steroids | HMDB0062657 | 0.11 [0.05, 0.18] | $5.8 \times 10^{-4}$ |
| androstenediol (3beta,17beta) disulfate (2) | Sulfated steroids | HMDB0240313 | 0.077 [0.025, 0.13] | 0.0045 |
| **Urea cycle; Arginine and Proline Metabolism** | | | | |
| N,N,N-trimethyl-alanylproline betaine (TMAP) | Amino acids, peptides, and analogues | HMDB0240365 | 0.076 [0.047, 0.11] | $1.2 \times 10^{-6}$ |
| arginine | Amino acids, peptides, and analogues | HMDB0000517 | 0.062 [0.025, 0.098] | 0.0012 |
| dimethylarginine (ADMA + SDMA) | Amino acids, peptides, and analogues | HMDB0003334 | -0.043 [-0.069, -0.017] | 0.0015 |
| ornithine | Amino acids, peptides, and analogues | HMDB0000214 | -0.05 [-0.098, -0.0025] | 0.041 |
| prolylhydroxyproline | Amino acids, peptides, and analogues | HMDB0006695 | -0.051 [-0.09, -0.012] | 0.011 |
| **Tyrosine Metabolism** | | | | |
| phenol sulfate | Arylsulfates | HMDB0060015 | 0.055 [0.012, 0.099] | 0.014 |
| 3-methoxytyrosine | Amino acids, peptides, and analogues | HMDB0001434 | -0.037 [-0.071, -0.0021] | 0.04 |
| 4-methoxyphenol sulfate | | | -0.065 [-0.12, -0.012] | 0.017 |
| N-formylphenylalanine | Amino acids, peptides, and analogues | HMDB0240317 | -0.067 [-0.12, -0.016] | 0.011 |
| N-acetyltyrosine | Amino acids, peptides, and analogues | HMDB0000866 | -0.071 [-0.12, -0.023] | 0.004 |
| **Chemical** | | | | |
| sulfate | Non-metal sulfates | HMDB0001448 | -0.032 [-0.062, -0.0016] | 0.041 |
| perfluorohexanesulfonate (PFHxS) | | HMDB0094700 | -0.069 [-0.13, -0.0067] | 0.032 |
| perfluorooctanesulfonate (PFOS) | | HMDB0059586 | -0.085 [-0.14, -0.032] | 0.0019 |
| perfluorooctanoate (PFOA) | | HMDB0059587 | -0.093 [-0.15, -0.04] | $8 \times 10^{-4}$ |
| 3-bromo-5-chloro-2,6-dihydroxybenzoic acid | Sulfated steroids | HMDB0242133 | -0.11 [-0.16, -0.067] | $3.6 \times 10^{-6}$ |
| **Fatty Acid, Monohydroxy** | | | | |
| 3-hydroxyhexanoate | Medium-chain hydroxy acids and derivatives | HMDB0061652 | -0.062 [-0.11, -0.018] | 0.0067 |
| 2-hydroxyheptanoate | | | -0.064 [-0.12, -0.011] | 0.019 |
| 3-hydroxysebacate | Medium-chain hydroxy acids and derivatives | HMDB0000350 | -0.069 [-0.12, -0.02] | 0.0068 |
| 3-hydroxydecanoate | Medium-chain hydroxy acids and derivatives | HMDB0002203 | -0.073 [-0.12, -0.025] | 0.0033 |
| 3-hydroxyoctanoate | Medium-chain hydroxy acids and derivatives | HMDB0001954 | -0.11 [-0.16, -0.059] | $4.7 \times 10^{-5}$ |
| **Histidine Metabolism** | | | | |
| N-acetylcarnosine | Amino acids (histidine metabolism) | HMDB0012881 | 0.065 [0.032, 0.098] | $1.5 \times 10^{-4}$ |
| imidazole lactate | Amino acids (histidine metabolism) | HMDB0002320 | 0.057 [0.016, 0.099] | 0.0072 |
| carnosine | Amino acids (histidine metabolism) | HMDB0000033 | -0.055 [-0.11, -0.0039] | 0.037 |
| N-acetylhistidine | Amino acids (histidine metabolism) | HMDB0032055 | -0.072 [-0.12, -0.029] | 0.0013 |
| **Corticosteroids** | | | | |
| tetrahydrocortisol glucuronide | | | 0.065 [0.023, 0.11] | 0.003 |
| cortisone | Hydroxysteroids | HMDB0002802 | 0.058 [0.018, 0.098] | 0.005 |
| tetrahydrocortisone glucuronide (5) | | | 0.05 [0.0052, 0.095] | 0.03 |
| cortolone glucuronide (1) | | | 0.047 [0.0057, 0.088] | 0.027 |
| **Phospholipid Metabolism** | | | | |
| trimethylamine N-oxide | Aminoxides | HMDB0000925 | 0.052 [0.0031, 0.1] | 0.039 |
| glycerophosphoethanolamine | Glycerophosphoethanolamines | HMDB0000114 | 0.044 [0.011, 0.078] | 0.0097 |
| glycerophosphorylcholine (GPC) | Glycerophosphocholines | HMDB0000086 | 0.032 [0.00089, 0.063] | 0.046 |
| phosphocholine | Quaternary ammonium salts | HMDB0001565 | -0.052 [-0.09, -0.015] | 0.0069 |
| **Glutamate Metabolism** | | | | |
| 2-pyrrolidinone | Pyrrolidones | HMDB0002039 | -0.036 [-0.07, -0.0028] | 0.036 |
| N-acetylglutamate | Amino acids, peptides, and analogues | HMDB0001138 | -0.049 [-0.095, -0.0028] | 0.039 |
| beta-citrylglutamate | Piperazines | HMDB0062295 | -0.08 [-0.13, -0.028] | 0.0031 |
| N-acetyl-aspartyl-glutamate (NAAG) | Amino acids, peptides, and analogues | HMDB0001067 | -0.11 [-0.15, -0.068] | $7 \times 10^{-7}$ |
| **Lysine Metabolism** | | | | |
| N6-acetyllysine | Amino acids, peptides, and analogues | HMDB0000206 | -0.046 [-0.084, -0.0086] | 0.017 |
| N-acetyl-2-aminoadipate | | | -0.048 [-0.088, -0.0073] | 0.022 |
| N6,N6-dimethyllysine | Amino acids, peptides, and analogues | HMDB0013287 | -0.048 [-0.088, -0.0087] | 0.018 |
| 2-aminoadipate | Amino acids, peptides, and analogues | HMDB0000510 | -0.05 [-0.095, -0.0057] | 0.029 |
| **Ascorbate and Aldarate Metabolism** | | | | |
| ascorbic acid 3-sulfate | Cinnamic acids | HMDB0125166 | 0.079 [0.027, 0.13] | 0.0033 |
| oxalate (ethanedioate) | Dicarboxylic acids and derivatives | HMDB0002329 | -0.044 [-0.085, -0.0026] | 0.039 |
| gulonate | Medium-chain hydroxy acids and derivatives | HMDB0003290 | -0.051 [-0.088, -0.013] | 0.009 |
| **Plasmalogen** | | | | |
| 1-(1-enyl-palmitoyl)-2-linoleoyl-GPC (P-16:0/18:2) | Glycerophosphocholines | HMDB0011211 | 0.061 [0.031, 0.092] | $1.2 \times 10^{-4}$ |
| 1-palmityl-2-linoleoyl-GPC (O-16:0/18:2) | Glycerophosphocholines | HMDB0011151 | 0.048 [0.012, 0.084] | 0.0092 |
| 1-(1-enyl-stearoyl)-2-linoleoyl-GPC (P-18:0/18:2) | Glycerophosphocholines | HMDB0011244 | 0.048 [0.014, 0.082] | 0.0067 |
| **Sterol** | | | | |
| beta-sitosterol | Stigmastanes and derivatives | HMDB0000852 | 0.059 [0.016, 0.1] | 0.0086 |
| 7-hydroxycholesterol (alpha or beta) | Cholestane steroids | HMDB0006119 | -0.079 [-0.13, -0.031] | 0.0016 |
| 4-cholesten-3-one | Cholestane steroids | HMDB0000921 | -0.085 [-0.13, -0.037] | $6.8 \times 10^{-4}$ |
| **Purine Metabolism, Adenine containing** | | | | |
| N6-carbamoylthreonyladenosine | | HMDB0041623 | -0.039 [-0.07, -0.0076] | 0.016 |
| N6-succinyladenosine | | HMDB0000912 | -0.05 [-0.086, -0.014] | 0.0077 |
| adenosine | | HMDB0000050 | -0.054 [-0.095, -0.014] | 0.0092 |
| **Pyrimidine Metabolism, Uracil containing** | | | | |
| 3-ureidopropionate | Ureas | HMDB0000026 | -0.047 [-0.081, -0.012] | 0.0086 |
| uridine | | HMDB0000296 | -0.061 [-0.097, -0.025] | 0.0012 |
| 2'-O-methyluridine | | HMDB0240328 | -0.071 [-0.11, -0.034] | $2.5 \times 10^{-4}$ |
| **Dihydrosphingomyelins** | | | | |
| sphingomyelin (d18:0/20:0, d16:0/22:0) | Phosphosphingolipids | HMDB0012090 | -0.052 [-0.091, -0.014] | 0.0087 |
| behenoyl dihydrosphingomyelin (d18:0/22:0) | Phosphosphingolipids | HMDB0012091 | -0.057 [-0.096, -0.017] | 0.0056 |
| sphingomyelin (d18:0/18:0, d19:0/17:0) | Phosphosphingolipids | HMDB0012087 | -0.065 [-0.1, -0.028] | $7.7 \times 10^{-4}$ |
| **Polyamine Metabolism** | | | | |
| N-acetyl-isoputreanine | Amino acids, peptides, and analogues | HMDB0242185 | -0.065 [-0.1, -0.028] | $6.6 \times 10^{-4}$ |
| spermidine | Amines | HMDB0001257 | -0.12 [-0.17, -0.058] | $1.1 \times 10^{-4}$ |
| spermine | Amines | HMDB0001256 | -0.12 [-0.18, -0.056] | $2.8 \times 10^{-4}$ |
| **Creatine Metabolism** | | | | |
| creatinine | Amino acids, peptides, and analogues | HMDB0000562 | 0.074 [0.046, 0.1] | $7.8 \times 10^{-7}$ |
| guanidinoacetate | Amino acids, peptides, and analogues | HMDB0000128 | 0.058 [0.023, 0.092] | 0.0012 |
| **Fructose, Mannose and Galactose Metabolism** | | | | |
| fructose | Carbohydrates and carbohydrate conjugates | HMDB0000660 | 0.067 [0.0066, 0.13] | 0.031 |
| galactonate | Medium-chain hydroxy acids and derivatives | HMDB0000565 | -0.1 [-0.17, -0.041] | 0.0015 |
| **Lactosylceramides (LCER)** | | | | |
| lactosyl-N-palmitoyl-sphingosine (d18:1/16:0) | Glycosphingolipids | HMDB0006750 | 0.043 [0.011, 0.075] | 0.0087 |
| lactosyl-N-nervonoyl-sphingosine (d18:1/24:1) | Glycosphingolipids | HMDB0004872 | 0.042 [7e-04, 0.084] | 0.048 |
| **Glycolysis, Gluconeogenesis, and Pyruvate Metabolism** | | | | |
| 1,5-anhydroglucitol (1,5-AG) | Carbohydrates and carbohydrate conjugates | HMDB0002712 | 0.041 [0.0051, 0.076] | 0.026 |
| pyruvate | Alpha-keto acids and derivatives | HMDB0000243 | -0.11 [-0.21, -0.003] | 0.046 |
| **Phenylalanine Metabolism** | | | | |
| 4-hydroxyphenylacetate | Benzoic acids and derivatives | HMDB0000020 | -0.039 [-0.077, -0.0018] | 0.042 |
| N-acetylphenylalanine | Amino acids, peptides, and analogues | HMDB0000512 | -0.096 [-0.14, -0.054] | $1.5 \times 10^{-5}$ |
| **Dihydroceramides** | | | | |
| N-palmitoyl-sphinganine (d18:0/16:0) | Ceramides | HMDB0011760 | -0.043 [-0.083, -0.0039] | 0.033 |
| N-stearoyl-sphinganine (d18:0/18:0) | Ceramides | HMDB0011761 | -0.045 [-0.082, -0.0081] | 0.018 |
| **Fatty Acid Metabolism (Acyl Glycine)** | | | | |
| picolinoylglycine | Amino acids, peptides, and analogues | HMDB0059766 | -0.045 [-0.085, -0.0045] | 0.031 |
| hexanoylglycine (C6) | Amino acids, peptides, and analogues | HMDB0000701 | -0.12 [-0.19, -0.057] | $3.3 \times 10^{-4}$ |
| **Glycine, Serine and Threonine Metabolism** | | | | |
| N-acetylserine | Amino acids, peptides, and analogues | HMDB0002931 | -0.045 [-0.076, -0.013] | 0.0065 |
| N-acetylglycine | Amino acids, peptides, and analogues | HMDB0000532 | -0.058 [-0.096, -0.02] | 0.0032 |
| **Pentose Metabolism** | | | | |
| ribonate | Carbohydrates and carbohydrate conjugates | HMDB0000867 | -0.045 [-0.087, -0.0032] | 0.037 |
| sedoheptulose | Carbohydrates and carbohydrate conjugates | HMDB0003219 | -0.083 [-0.13, -0.035] | $8.3 \times 10^{-4}$ |
| **Nicotinate and Nicotinamide Metabolism** | | | | |
| quinolinate | Pyridinecarboxylic acids and derivatives | HMDB0000232 | -0.046 [-0.084, -0.0091] | 0.016 |
| nicotinamide | Pyridinecarboxylic acids and derivatives | HMDB0001406 | -0.086 [-0.14, -0.03] | 0.0029 |
| **Purine Metabolism, Guanine containing** | | | | |
| N2,N2-dimethylguanosine | | HMDB0004824 | -0.048 [-0.084, -0.011] | 0.012 |
| 7-methylguanine | Purines and purine derivatives | HMDB0000897 | -0.06 [-0.089, -0.03] | $1.3 \times 10^{-4}$ |
| **Endocannabinoid** | | | | |
| N-palmitoylserine | Arylsulfates | HMDB0170765 | -0.049 [-0.095, -0.0038] | 0.036 |
| hexanoyltaurine | | | -0.099 [-0.17, -0.026] | 0.0086 |
| **Glutathione Metabolism** | | | | |
| 5-oxoproline | Amino acids, peptides, and analogues | HMDB0000267 | -0.051 [-0.091, -0.011] | 0.014 |
| cysteinylglycine disulfide | Amino acids, peptides, and analogues | HMDB0000709 | -0.073 [-0.12, -0.027] | 0.0021 |
| **Aminosugar Metabolism** | | | | |
| N-acetylneuraminate | Carbohydrates and carbohydrate conjugates | HMDB0000230 | -0.054 [-0.09, -0.018] | 0.0038 |
| N-acetylglucosamine/N-acetylgalactosamine | Carbohydrates and carbohydrate conjugates | HMDB0000212 | -0.066 [-0.1, -0.03] | $5.3 \times 10^{-4}$ |
| **Purine Metabolism, (Hypo)Xanthine/Inosine containing** | | | | |
| xanthine | Purines and purine derivatives | HMDB0000292 | -0.066 [-0.12, -0.016] | 0.01 |
| allantoin | Imidazoles | HMDB0000462 | -0.1 [-0.15, -0.049] | $2 \times 10^{-4}$ |
| **Modified Peptides** | | | | |
| N,N-dimethyl-pro-pro | | | -0.067 [-0.1, -0.03] | $4.5 \times 10^{-4}$ |
| pyroglutamylvaline | Fatty acids and conjugates | HMDB0245154 | -0.11 [-0.17, -0.046] | $7.1 \times 10^{-4}$ |
| **Pyrimidine Metabolism, Orotate containing** | | | | |
| orotidine | | HMDB0000788 | -0.081 [-0.12, -0.039] | $2.8 \times 10^{-4}$ |
| orotate | Pyrimidines and pyrimidine derivatives | HMDB0000226 | -0.083 [-0.14, -0.027] | 0.0044 |
| **Dipeptide Derivative** | | | | |
| isoleucylhydroxyproline | Amino acids, peptides, and analogues | HMDB0028908 | -0.083 [-0.13, -0.039] | $2.9 \times 10^{-4}$ |
| leucylhydroxyproline | Amino acids, peptides, and analogues | HMDB0028930 | -0.12 [-0.19, -0.044] | 0.0024 |
| **Tocopherol Metabolism** | | | | |
| alpha-tocopherol | Quinone and hydroquinone lipids | HMDB0001893 | 0.11 [0.031, 0.19] | 0.007 |
| **Vitamin A Metabolism** | | | | |
| retinol (vitamin A) | Retinoids | HMDB0000305 | 0.093 [0.019, 0.17] | 0.015 |
| **Alanine and Aspartate Metabolism** | | | | |
| N,N-dimethylalanine | | | 0.091 [0.044, 0.14] | $2.4 \times 10^{-4}$ |
| **Secondary Bile Acid Metabolism** | | | | |
| glycolithocholate sulfate | Bile acids, alcohols and derivatives | HMDB0002639 | 0.07 [0.0082, 0.13] | 0.028 |
| **Drug - Topical Agents** | | | | |
| salicylate | Benzoic acids and derivatives | HMDB0001895 | 0.065 [0.0031, 0.13] | 0.041 |
| **Eicosanoid** | | | | |
| leukotriene B4 | Eicosanoids | HMDB0001085 | 0.057 [0.021, 0.092] | 0.0019 |
| **Monoacylglycerol** | | | | |
| 1-palmitoleoylglycerol (16:1) | Monoradylglycerols | HMDB0011565 | 0.052 [0.0032, 0.1] | 0.039 |
| **Phosphatidylcholine (PC)** | | | | |
| 1-stearoyl-2-(hydroxylinoleoyl)-GPC (18:0/18:2(OH)) | | | 0.049 [0.015, 0.082] | 0.0052 |
| **Progestin Steroids** | | | | |
| 5alpha-pregnan-3beta,20beta-diol monosulfate (1) | Sulfated steroids | HMDB0240580 | 0.048 [0.011, 0.085] | 0.013 |
| **Sphingomyelins** | | | | |
| lignoceroyl sphingomyelin (d18:1/24:0) | Phosphosphingolipids | HMDB0011697 | 0.036 [0.0046, 0.068] | 0.027 |
| **Long Chain Monounsaturated Fatty Acid** | | | | |
| 10-nonadecenoate (19:1n9) | Fatty acids and conjugates | HMDB0013622 | -0.056 [-0.11, -0.003] | 0.04 |
| **Guanidino and Acetamido Metabolism** | | | | |
| guanidinosuccinate | Amino acids, peptides, and analogues | HMDB0003157 | -0.056 [-0.097, -0.015] | 0.0079 |
| **Fatty Acid Metabolism (Acyl Carnitine, Long Chain Saturated)** | | | | |
| margaroylcarnitine (C17) | Fatty acid esters | HMDB0006210 | -0.057 [-0.11, -0.0035] | 0.039 |
| **Fatty Acid Synthesis** | | | | |
| malonate | Dicarboxylic acids and derivatives | HMDB0000691 | -0.058 [-0.095, -0.02] | 0.0029 |
| **Vitamin B6 Metabolism** | | | | |
| pyridoxate | Pyridinecarboxylic acids and derivatives | HMDB0000017 | -0.063 [-0.12, -0.0012] | 0.048 |
| **Short Chain Fatty Acid** | | | | |
| valerate (5:0) | Fatty acids and conjugates | HMDB0000892 | -0.067 [-0.11, -0.02] | 0.0063 |
| **Pyrimidine Metabolism, Thymine containing** | | | | |
| 3-aminoisobutyrate | Amino acids, peptides, and analogues | HMDB0002166 | -0.073 [-0.12, -0.03] | 0.001 |
| **Partially Characterized Molecules** | | | | |
| branched-chain, straight-chain, or cyclopropyl 10:1 fatty acid (1) | | | -0.081 [-0.14, -0.018] | 0.013 |
| **TCA Cycle** | | | | |
| alpha-ketoglutarate | Gamma-keto acids and derivatives | HMDB0000208 | -0.091 [-0.16, -0.018] | 0.016 |
| **Fatty Acid Metabolism (Acyl Glutamine)** | | | | |
| hexanoylglutamine | Amino acids, peptides, and analogues | HMDB0242182 | -0.093 [-0.14, -0.049] | $6.9 \times 10^{-5}$ |
| **Fatty Acid, Oxidized** | | | | |
| 4-hydroxynonenal | Fatty alcohols | HMDB0004362 | -0.1 [-0.17, -0.033] | 0.0045 |
| **Ketone Bodies** | | | | |
| 3-hydroxybutyrate (BHBA) | Beta hydroxy acids and derivatives | HMDB0000442 | -0.1 [-0.16, -0.041] | 0.0013 |
| **Oxidative Phosphorylation** | | | | |
| phosphate | Non-metal phosphates | HMDB0001429 | -0.11 [-0.17, -0.053] | $3.2 \times 10^{-4}$ |

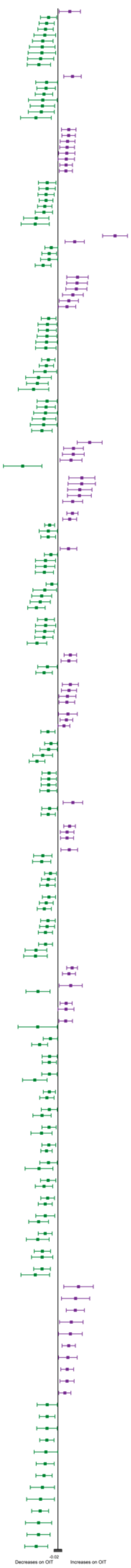

Supplemental Figure 2. Biological pathways significantly associated with change over time on OIT. Pathways enriched in both *DEVIL* and *PNOIT* are indicated in orange and scaled by the percent of matching metabolites.

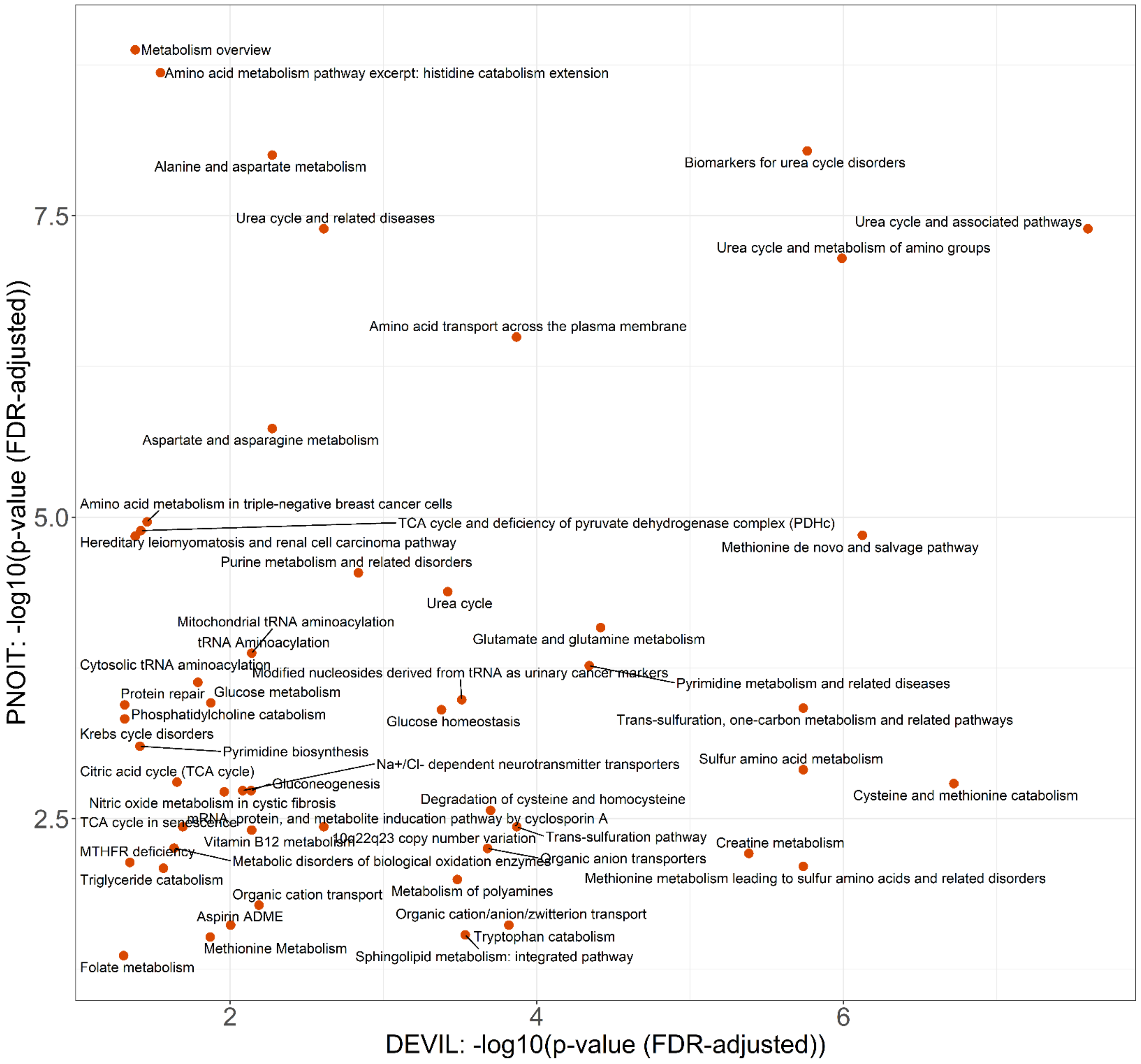

Supplemental Table 2. Summary of chemical subclass enrichment of metabolites significantly associated with time on OIT in both *DEVIL* and *PNOIT*.

| **Subclass** | **P-val (FDR adjusted) (*DEVIL*)** | **Enrichment Ratio (*DEVIL*)** | **P-val (FDR adjusted) (*PNOIT*)** | **Enrichment Ratio (*PNOIT*)** | **Enrichment Ratio Mean (*DEVIL*& *PNOIT*)** |
|---|---|---|---|---|---|
| Amino acids, peptides, and analogues | 6.1E-45 | 13.3 | 2.1E-05 | 4.7 | 9.0 |
| Medium-chain hydroxy acids and derivatives | 4.4E-13 | 102.5 | | | 102.5 |

| | | | | | |
|---|---|---|---|---|---|
| Sulfated steroids | 4.5E-10 | 107.3 | | | 107.3 |
| Arylsulfates | 4.4E-08 | 48.6 | | | 48.6 |
| Fatty acids and conjugates | 5.6E-08 | 9.7 | 1.2E-05 | 9.9 | 9.8 |
| Phosphosphingolipids | 3.8E-05 | 37.3 | | | 37.3 |
| Dicarboxylic acids and derivatives | 0.0013 | 26.8 | 0.09860 | 13.7 | 20.3 |
| Pyridinecarboxylic acids and derivatives | 0.0013 | 26.5 | | | 26.5 |
| Short-chain keto acids and derivatives | 0.0036 | 53.6 | | | 53.6 |
| Glycerophosphocholines | 0.0058 | 4.4 | 1.0E-13 | 15.5 | 10.0 |
| Beta hydroxy acids and derivatives | 0.0064 | 36.5 | 0.05267 | 28.0 | 32.3 |
| Ceramides | 0.0090 | 29.1 | | | 29.1 |
| Non-metal sulfates | 0.0090 | 429.1 | | | 429.1 |
| Aminoxides | 0.0125 | 286.1 | | | 286.1 |
| Non-metal phosphates | 0.0155 | 214.6 | | | 214.6 |
| Short-chain hydroxy acids and derivatives | 0.0171 | 171.7 | | | 171.7 |
| Sulfinic acids | 0.0171 | 171.7 | 0.00725 | 263.1 | 217.4 |
| Hybrid peptides | 0.0276 | 13.1 | | | 13.1 |
| Imidazoles | 0.0276 | 12.5 | | | 12.5 |
| Cinnamic acids | 0.0276 | 85.8 | | | 85.8 |
| Piperidinones | 0.0276 | 85.8 | | | 85.8 |
| Alpha-keto acids and derivatives | 0.0316 | 71.5 | | | 71.5 |
| Cholestane steroids | 0.0323 | 10.9 | | | 10.9 |
| Carbohydrates and carbohydrate conjugates | 0.0391 | 2.9 | | | 2.9 |
| Gamma-keto acids and derivatives | 0.0529 | 37.3 | 0.02801 | 57.2 | 47.3 |
| Pyrrolidones | 0.0683 | 27.7 | | | 27.7 |
| Purines and purine derivatives | 0.0731 | 6.4 | 0.20665 | 4.9 | 5.7 |
| Ureas | 0.0774 | 22.6 | | | 22.6 |
| Medium-chain keto acids and derivatives | 0.0870 | 19.1 | | | 19.1 |
| 1-hydroxy-2-unsubstituted benzenoids | 0.0870 | 18.7 | | | 18.7 |
| Quaternary ammonium salts | 0.10 | 15.3 | | | 15.3 |
| Retinoids | 0.11 | 13.6 | | | 13.6 |
| Amines | 0.11 | 4.6 | 0.00637 | 10.5 | 7.6 |
| Quinolones and derivatives | 0.12 | 11.9 | | | 11.9 |
| Quinoline carboxylic acids | 0.12 | 11.8 | | | 11.8 |
| Quinone and hydroquinone lipids | 0.15 | 8.6 | | | 8.6 |
| Glycosphingolipids | 0.16 | 3.4 | | | 3.4 |
| Monoradylglycerols | 0.16 | 7.5 | | | 7.5 |
| Stigmastanes and derivatives | 0.18 | 6.5 | | | 6.5 |
| Hydroxysteroids | 0.19 | 6.0 | | | 6.0 |
| Benzoic acids and derivatives | 0.22 | 2.6 | 5.3E-05 | 11.9 | 7.3 |
| Piperazines | 0.23 | 4.8 | | | 4.8 |
| Pyrimidines and pyrimidine derivatives | 0.30 | 3.4 | | | 3.4 |
| Bile acids, alcohols and derivatives | 0.32 | 3.0 | | | 3.0 |
| Carbonyl compounds | 0.38 | 1.7 | 0.20665 | 2.5 | 2.1 |
| Eicosanoids | 0.47 | 1.8 | 1.8E-20 | 43.8 | 22.8 |
| Diterpenoids | 0.48 | 1.7 | | | 1.7 |
| Fatty alcohols | 0.48 | 1.6 | | | 1.6 |

| | | | | | |
|---|---|---|---|---|---|
| Glycerophosphoethanolamines | 0.71 | 0.8 | 0.00034 | 4.3 | 2.5 |
| Fatty acid esters | 0.86 | 0.5 | 0.17723 | 2.4 | 1.4 |
| Steroid esters | | | 6.0E-05 | 26.7 | 26.7 |
| Tricarboxylic acids and derivatives | | | 0.00013 | 45.9 | 45.9 |
| Linoleic acids and derivatives | | | 0.00044 | 14.4 | 14.4 |
| Hydroxyindoles | | | 0.02773 | 62.6 | 62.6 |
| Diradylglycerols | | | 0.20665 | 1.9 | 1.9 |
| Glycerophosphoserines | | | 0.53592 | 1.4 | 1.4 |
| Triacylglycerols | | | 1.00000 | 0.3 | 0.3 |

Supplemental Table 3. Summary of biological pathways enriched based on metabolites significantly associated with time on OIT in both *DEVIL* and *PNOIT*.

| **Biological Pathway** | **Pathway ID** | **Pathway Source** | **P-val (FDR adjusted) (DEVIL)** | **% Matched Metabolites (*DEVIL*)** | **P-val (FDR adjusted) (PNOIT)** | **% Matched Metaboites (*PNOIT*)** |
|---|---|---|---|---|---|---|
| Urea cycle and related diseases | WP4571 | wiki | 2.46E-03 | 21.4 | 4.06E-08 | 35.7 |
| Biomarkers for urea cycle disorders | WP4583 | wiki | 1.73E-06 | 27.3 | 9.19E-09 | 27.3 |
| Amino acid metabolism pathway excerpt: histidine catabolism extension | WP4661 | wiki | 2.85E-02 | 11.8 | 2.06E-09 | 35.3 |
| Urea cycle and associated pathways | WP4595 | wiki | 2.55E-08 | 26.7 | 4.06E-08 | 20.0 |
| Creatine metabolism | R-HSA-71288 | reactome | 4.15E-06 | 33.3 | 6.13E-03 | 13.3 |
| Nitric oxide metabolism in cystic fibrosis | WP4947 | wiki | 1.09E-02 | 22.2 | 1.89E-03 | 22.2 |
| Trans-sulfuration pathway | WP2333 | wiki | 1.35E-04 | 28.6 | 3.69E-03 | 14.3 |
| Alanine and aspartate metabolism | WP106 | wiki | 5.32E-03 | 13.0 | 9.92E-09 | 26.1 |
| Protein repair | R-HSA-5676934 | reactome | 1.63E-02 | 15.4 | 2.34E-04 | 23.1 |
| Urea cycle and metabolism of amino groups | WP497 | wiki | 1.03E-06 | 20.6 | 7.14E-08 | 17.6 |
| Modified nucleosides derived from tRNA as urinary cancer markers | WP4485 | wiki | 3.09E-04 | 21.1 | 3.26E-04 | 15.8 |
| Organic anion transporters | R-HSA-428643 | reactome | 2.10E-04 | 22.2 | 5.56E-03 | 14.3 |
| Urea cycle | R-HSA-70635 | reactome | 3.81E-04 | 18.2 | 4.14E-05 | 18.2 |
| 10q22q23 copy number variation | WP5402 | wiki | 2.46E-03 | 21.4 | 3.69E-03 | 14.3 |
| Glutamate and glutamine metabolism | R-HSA-8964539 | reactome | 3.84E-05 | 20.0 | 8.22E-05 | 14.8 |

| | | | | | | |
|---|---|---|---|---|---|---|
| Methionine metabolism leading to sulfur amino acids and related disorders | WP4292 | wiki | 1.83E-06 | 26.1 | 7.85E-03 | 8.7 |
| Glucose homeostasis | WP661 | wiki | 4.19E-04 | 19.0 | 3.95E-04 | 14.3 |
| Aspartate and asparagine metabolism | R-HSA-8963693 | reactome | 5.32E-03 | 12.0 | 1.84E-06 | 20.8 |
| Amino acid transport across the plasma membrane | R-HSA-352230 | reactome | 1.36E-04 | 14.7 | 3.21E-07 | 17.6 |
| Amino acid metabolism in triple-negative breast cancer cells | WP5213 | wiki | 3.49E-02 | 10.5 | 1.09E-05 | 21.1 |
| TCA cycle and deficiency of pyruvate dehydrogenase complex (PDHc) | WP2453 | wiki | 3.83E-02 | 10.0 | 1.29E-05 | 20.0 |
| Metabolism of polyamines | R-HSA-351202 | reactome | 3.30E-04 | 19.0 | 1.01E-02 | 9.5 |
| TCA cycle in senescence | WP5050 | wiki | 2.04E-02 | 14.3 | 3.69E-03 | 14.3 |
| Hereditary leiomyomatosis and renal cell carcinoma pathway | WP4206 | wiki | 4.16E-02 | 9.5 | 1.44E-05 | 19.0 |
| mRNA, protein, and metabolite inducation pathway by cyclosporin A | WP3953 | wiki | 2.04E-02 | 14.3 | 3.69E-03 | 14.3 |
| Cysteine and methionine catabolism | WP4504 | wiki | 1.91E-07 | 20.5 | 1.62E-03 | 7.7 |
| TCA cycle (aka Krebs or citric acid cycle) | WP78 | wiki | 5.66E-02 | 8.0 | 6.98E-07 | 20.0 |
| Citric Acid Cycle | map00020 | kegg | 1.66E-01 | 7.7 | 6.37E-04 | 19.2 |
| Methionine de novo and salvage pathway | WP3580 | wiki | 7.54E-07 | 16.3 | 1.41E-05 | 10.2 |
| Phosphatidylcholine catabolism | WP4195 | wiki | 4.88E-02 | 8.7 | 3.59E-04 | 15.0 |
| Nitrogen metabolism | map00910 | kegg | 1.57E-01 | 8.0 | 3.92E-03 | 15.4 |
| Pyrimidine biosynthesis | R-HSA-500753 | reactome | 3.88E-02 | 9.1 | 7.92E-04 | 13.6 |
| Pyrimidine metabolism and related diseases | WP4225 | wiki | 4.56E-05 | 13.6 | 1.70E-04 | 9.1 |
| Triglyceride catabolism | R-HSA-163560 | reactome | 2.73E-02 | 11.1 | 8.12E-03 | 11.1 |
| Krebs cycle disorders | WP4236 | wiki | 4.88E-02 | 8.7 | 4.70E-04 | 13.0 |
| Degradation of cysteine and homocysteine | R-HSA-1614558 | reactome | 2.00E-04 | 13.2 | 2.70E-03 | 7.9 |

| | | | | | | |
|---|---|---|---|---|---|---|
| Trans-sulfuration, one-carbon metabolism and related pathways | WP2525 | wiki | 1.83E-06 | 13.3 | 3.82E-04 | 6.9 |
| Organic cation/anion/zwitterion transport | R-HSA-549132 | reactome | 1.52E-04 | 14.3 | 2.41E-02 | 5.7 |
| Tryptophan catabolism | R-HSA-71240 | reactome | 1.52E-04 | 14.3 | 2.41E-02 | 5.7 |
| Arginine and proline metabolism | map00330 | kegg | 5.15E-02 | 9.8 | 6.71E-03 | 9.8 |
| Methionine Metabolism | map00270 | kegg | 1.35E-02 | 11.8 | 3.02E-02 | 7.8 |
| Gluconeogenesis | R-HSA-70263 | reactome | 7.31E-03 | 10.0 | 1.85E-03 | 9.4 |
| Na+/Cl- dependent neurotransmitter transporters | R-HSA-442660 | reactome | 8.29E-03 | 9.4 | 1.85E-03 | 9.4 |
| Metabolic reprogramming in colon cancer | WP4290 | wiki | 6.25E-02 | 5.3 | 4.78E-06 | 13.2 |
| MTHFR deficiency | WP4288 | wiki | 4.51E-02 | 9.1 | 7.28E-03 | 9.1 |
| Sulfur amino acid metabolism | R-HSA-1614635 | reactome | 1.83E-06 | 12.1 | 1.24E-03 | 6.1 |
| Organic cation transport | R-HSA-549127 | reactome | 6.49E-03 | 10.7 | 1.64E-02 | 7.1 |
| beta-Alanine metabolism | map00410 | kegg | 6.73E-02 | 10.3 | 1.13E-01 | 6.9 |
| Purine metabolism and related disorders | WP4224 | wiki | 1.46E-03 | 8.5 | 2.89E-05 | 8.5 |
| Glucose metabolism | R-HSA-70326 | reactome | 1.34E-02 | 7.3 | 3.46E-04 | 9.3 |
| Alanine, aspartate and glutamate metabolism | map00250 | kegg | 6.21E-02 | 8.2 | 1.21E-02 | 8.2 |
| NAD biosynthesis II from tryptophan | WP2485 | wiki | 5.66E-02 | 8.0 | 8.38E-03 | 8.3 |
| Citric acid cycle (TCA cycle) | R-HSA-71403 | reactome | 2.23E-02 | 5.8 | 1.57E-03 | 10.3 |
| Phospholipid Biosynthesis | map00564 | kegg | 1.57E-01 | 8.0 | 9.52E-02 | 8.0 |
| Metabolic pathways of fibroblasts | WP5312 | wiki | 5.66E-02 | 8.0 | 8.76E-03 | 8.0 |
| Phenylalanine and tyrosine metabolism | R-HSA-8963691 | reactome | 6.21E-02 | 6.1 | 1.74E-03 | 9.7 |
| Aspirin ADME | R-HSA-9749641 | reactome | 9.94E-03 | 8.6 | 2.41E-02 | 5.7 |
| Triglyceride metabolism | R-HSA-8979227 | reactome | 5.95E-02 | 7.1 | 1.64E-02 | 7.1 |
| Nicotinate and nicotinamide metabolism | map00760 | kegg | 1.95E-01 | 6.9 | 1.13E-01 | 6.9 |
| Cytosolic tRNA aminoacylation | R-HSA-379716 | reactome | 7.24E-03 | 6.1 | 1.34E-04 | 7.6 |

| | | | | | | |
|---|---|---|---|---|---|---|
| tRNA Aminoacylation | R-HSA-379724 | reactome | 7.24E-03 | 6.1 | 1.34E-04 | 7.6 |
| Mitochondrial tRNA aminoacylation | R-HSA-379726 | reactome | 7.24E-03 | 6.1 | 1.34E-04 | 7.6 |
| Tryptophan metabolism | map00380 | kegg | 6.21E-02 | 9.3 | 2.69E-01 | 3.7 |
| Sphingolipid metabolism: integrated pathway | WP4726 | wiki | 2.93E-04 | 9.0 | 2.90E-02 | 3.9 |
| Amino Sugar Metabolism | map00520 | kegg | 1.03E-01 | 7.7 | 1.69E-01 | 5.1 |
| Metabolism of fat-soluble vitamins | R-HSA-6806667 | reactome | 6.45E-02 | 4.3 | 4.24E-04 | 8.5 |
| Ferroptosis | WP4313 | wiki | 6.25E-02 | 5.0 | 1.62E-03 | 7.7 |
| Nucleotide biosynthesis | R-HSA-8956320 | reactome | 6.54E-02 | 4.2 | 4.53E-04 | 8.3 |
| 16p11.2 proximal deletion syndrome | WP4949 | wiki | 6.44E-02 | 4.8 | 1.71E-03 | 7.5 |
| Metabolism overview | WP3602 | wiki | 4.16E-02 | 3.4 | 1.33E-09 | 8.5 |
| Fatty acyl-CoA biosynthesis | R-HSA-75105 | reactome | 6.25E-02 | 4.9 | 3.69E-03 | 6.8 |
| Vitamin B12 metabolism | WP1533 | wiki | 7.24E-03 | 6.6 | 3.92E-03 | 4.9 |
| Digestion | R-HSA-8935690 | reactome | 6.30E-02 | 4.5 | 3.69E-03 | 6.8 |
| Metabolic disorders of biological oxidation enzymes | R-HSA-5579029 | reactome | 2.32E-02 | 5.7 | 5.54E-03 | 5.7 |
| Digestion and absorption | R-HSA-8963743 | reactome | 6.44E-02 | 4.3 | 3.87E-03 | 6.5 |
| Neurotransmitter release cycle | R-HSA-112310 | reactome | 6.45E-02 | 4.3 | 3.97E-03 | 6.4 |
| Glyoxylate metabolism and glycine degradation | R-HSA-389661 | reactome | 6.21E-02 | 5.4 | 3.13E-02 | 4.9 |
| Nicotinate metabolism | R-HSA-196807 | reactome | 5.63E-03 | 6.8 | 5.35E-02 | 3.3 |
| Folate metabolism | WP176 | wiki | 4.95E-02 | 4.5 | 4.31E-02 | 3.0 |
| Tyrosine metabolism | map00350 | kegg | 5.77E-01 | 2.9 | 1.43E-01 | 4.3 |
| Class C/3 (Metabotropic glutamate/pheromone receptors) | R-HSA-420499 | reactome | 7.28E-02 | 3.5 | 5.02E-02 | 3.5 |
| Peroxisomal lipid metabolism | R-HSA-390918 | reactome | 7.85E-02 | 3.3 | 5.36E-02 | 3.2 |
| Synthesis of substrates in N-glycan biosythesis | R-HSA-446219 | reactome | 1.05E-01 | 2.6 | 6.66E-02 | 2.6 |

Supplemental Table 4. Summary of chemical subclass enrichment of metabolites significantly associated with time on OIT.

| Subclass | P-val (FDR adjusted) | Enrichment Ratio | Percent Hits (%) |
|---|---|---|---|
| Amino acids, peptides, and analogues | 6.10E-45 | 13.3 | 1.6 |
| Medium-chain hydroxy acids and derivatives | 4.39E-13 | 102.5 | 11.9 |
| Sulfated steroids | 4.48E-10 | 107.3 | 12.5 |
| Arylsulfates | 4.42E-08 | 48.6 | 5.7 |
| Fatty acids and conjugates | 5.63E-08 | 9.7 | 1.1 |
| Phosphosphingolipids | 3.82E-05 | 37.3 | 4.3 |
| Dicarboxylic acids and derivatives | 0.0013 | 26.8 | 3.1 |
| Pyridinecarboxylic acids and derivatives | 0.0013 | 26.5 | 3.1 |
| Short-chain keto acids and derivatives | 0.0036 | 53.6 | 6.3 |
| Glycerophosphocholines | 0.0058 | 4.4 | 0.5 |
| Beta hydroxy acids and derivatives | 0.0064 | 36.5 | 4.3 |
| Ceramides | 0.0090 | 29.1 | 3.4 |
| Non-metal sulfates | 0.0090 | 429.1 | 50.0 |
| Aminoxides | 0.0125 | 286.1 | 33.3 |
| Non-metal phosphates | 0.0155 | 214.6 | 25.0 |
| Short-chain hydroxy acids and derivatives | 0.0171 | 171.7 | 20.0 |
| Sulfinic acids | 0.0171 | 171.7 | 20.0 |
| Hybrid peptides | 0.0276 | 13.1 | 1.5 |
| Imidazoles | 0.0276 | 12.5 | 1.5 |
| Cinnamic acids | 0.0276 | 85.8 | 10.0 |
| Piperidinones | 0.0276 | 85.8 | 10.0 |
| Alpha-keto acids and derivatives | 0.0316 | 71.5 | 8.3 |
| Cholestane steroids | 0.0323 | 10.9 | 1.3 |
| Carbohydrates and carbohydrate conjugates | 0.0391 | 2.9 | 0.3 |
| Gamma-keto acids and derivatives | 0.0529 | 37.3 | 4.3 |
| Pyrrolidones | 0.0683 | 27.7 | 3.2 |
| Purines and purine derivatives | 0.0731 | 6.4 | 0.7 |
| Ureas | 0.0774 | 22.6 | 2.6 |
| Medium-chain keto acids and derivatives | 0.0870 | 19.1 | 2.2 |
| 1-hydroxy-2-unsubstituted benzenoids | 0.0870 | 18.7 | 2.2 |
| Quaternary ammonium salts | 0.10 | 15.3 | 1.8 |
| Retinoids | 0.11 | 13.6 | 1.6 |
| Amines | 0.11 | 4.6 | 0.5 |
| Quinolones and derivatives | 0.12 | 11.9 | 1.4 |
| Quinoline carboxylic acids | 0.12 | 11.8 | 1.4 |

| | | | |
|---|---|---|---|
| Quinone and hydroquinone lipids | 0.15 | 8.6 | 1.0 |
| Glycosphingolipids | 0.16 | 3.4 | 0.4 |
| Monoradylglycerols | 0.16 | 7.5 | 0.9 |
| Stigmastanes and derivatives | 0.18 | 6.5 | 0.8 |
| Hydroxysteroids | 0.19 | 6.0 | 0.7 |
| Benzoic acids and derivatives | 0.22 | 2.6 | 0.3 |
| Piperazines | 0.23 | 4.8 | 0.6 |
| Pyrimidines and pyrimidine derivatives | 0.30 | 3.4 | 0.4 |
| Bile acids, alcohols and derivatives | 0.32 | 3.0 | 0.4 |
| Carbonyl compounds | 0.38 | 1.7 | 0.2 |
| Eicosanoids | 0.47 | 1.8 | 0.2 |
| Diterpenoids | 0.48 | 1.7 | 0.2 |
| Fatty alcohols | 0.48 | 1.6 | 0.2 |
| Glycerophosphoethanolamines | 0.71 | 0.8 | 0.1 |
| Fatty acid esters | 0.86 | 0.5 | 0.1 |

Supplemental Figure 3. Biological pathways enrichment of metabolites associated with change over time during OIT by RAMP-based biological pathway analysis. The percent of matched metabolites in pathways plotted by the FDR-adjusted enrichment p-value. Dark pink represents subclasses or pathways linked to food allergy in prior publications.

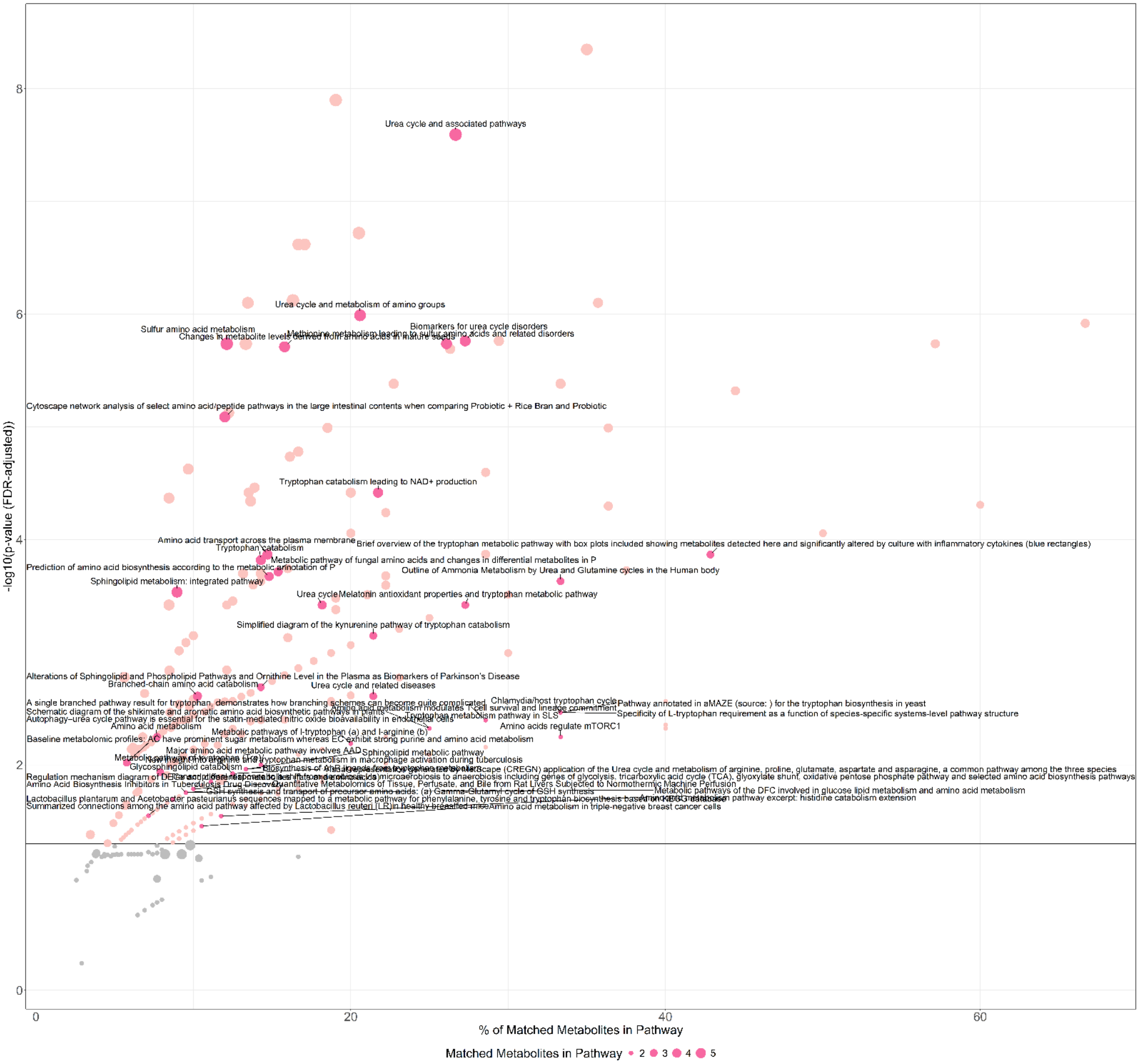

Supplemental Table 5. Summary of biological pathways enriched based on metabolites significantly associated with time on OIT.

| Biological Pathway | Pathway ID | Pathway Source | P-val (FDR adjusted) | % Matched Metabolites |
|---|---|---|---|---|
| Urea cycle and associated pathways | WP4595 | wiki | 2.55E-08 | 26.7 |
| Urea cycle and metabolism of amino groups | WP497 | wiki | 1.03E-06 | 20.6 |
| Biomarkers for urea cycle disorders | WP4583 | wiki | 1.73E-06 | 27.3 |
| Sulfur amino acid metabolism | R-HSA-1614635 | reactome | 1.83E-06 | 12.1 |
| Methionine metabolism leading to sulfur amino acids and related disorders | WP4292 | wiki | 1.83E-06 | 26.1 |
| Changes in metabolite levels derived from amino acids in mature seeds | PMC3901229__F4 | pfocr | 1.95E-06 | 15.8 |
| Cytoscape network analysis of select amino acid/peptide pathways in the large intestinal contents when comparing Probiotic + Rice Bran and Probiotic | PMC5399067__F5 | pfocr | 8.18E-06 | 12.0 |
| Tryptophan catabolism leading to NAD+ production | WP4210 | wiki | 3.84E-05 | 21.7 |
| Amino acid transport across the plasma membrane | R-HSA-352230 | reactome | 0.0001 | 14.7 |
| Brief overview of the tryptophan metabolic pathway with box plots included showing metabolites detected here and significantly altered by culture with inflammatory cytokines (blue rectangles) | PMC5557342__F5 | pfocr | 0.0001 | 42.9 |
| Tryptophan catabolism | R-HSA-71240 | reactome | 0.0002 | 14.3 |
| Metabolic pathway of fungal amino acids and changes in differential metabolites in P | PMC6990131__F5 | pfocr | 0.0002 | 15.4 |
| Prediction of amino acid biosynthesis according to the metabolic annotation of P | PMC6598237__F4 | pfocr | 0.0002 | 14.8 |
| Outline of Ammonia Metabolism by Urea and Glutamine cycles in the Human body | PMC10135863__F1 | pfocr | 0.0002 | 33.3 |
| Sphingolipid metabolism: integrated pathway | WP4726 | wiki | 0.0003 | 9.0 |
| Urea cycle | R-HSA-70635 | reactome | 0.0004 | 18.2 |
| Melatonin antioxidant properties and tryptophan metabolic pathway | PMC10216109__F2 | pfocr | 0.0004 | 27.3 |
| Simplified diagram of the kynurenine pathway of tryptophan catabolism | PMC7047773__F1 | pfocr | 0.0007 | 21.4 |
| Alterations of Sphingolipid and Phospholipid Pathways and Ornithine Level in the Plasma as Biomarkers of Parkinsonâ€™s Disease | PMC8834036__F5 | pfocr | 0.0020 | 14.3 |
| Branched-chain amino acid catabolism | R-HSA-70895 | reactome | 0.0025 | 10.3 |
| Urea cycle and related diseases | WP4571 | wiki | 0.0025 | 21.4 |
| Pathway annotated in aMAZE (source: ) for the tryptophan biosynthesis in yeast | PMC1160198__F2 | pfocr | 0.0034 | 33.3 |

| | | | | |
|---|---|---|---|---|
| Chlamydia/host tryptophan cycle | PMC126876__F2 | pfocr | 0.0034 | 33.3 |
| Specificity of L-tryptophan requirement as a function of species-specific systems-level pathway structure | PMC7084086__F4 | pfocr | 0.0034 | 33.3 |
| Amino acid metabolism modulates T cell survival and lineage commitment | PMC11061161__F3 | pfocr | 0.0034 | 33.3 |
| A single branched pathway result for tryptophan, demonstrates how branching schemes can become quite complicated | PMC2881407__F4 | pfocr | 0.0040 | 28.6 |
| Tryptophan metabolism pathway in SLS | PMC10301067__F4 | pfocr | 0.0040 | 28.6 |
| Schematic diagram of the shikimate and aromatic amino acid biosynthetic pathways in plants | PMC5597090__F2 | pfocr | 0.0047 | 25.0 |
| Autophagyâ€“urea cycle pathway is essential for the statin-mediated nitric oxide bioavailability in endothelial cells | PMC10629920__F8 | pfocr | 0.0047 | 25.0 |
| Amino acids regulate mTORC1 | R-HSA-9639288 | reactome | 0.0056 | 33.3 |
| Amino acid metabolism | PMC10497558__F1 | pfocr | 0.0056 | 7.7 |
| Metabolic pathways of l-tryptophan (a) and l-arginine (b) | PMC7563518__F1 | pfocr | 0.0065 | 20.0 |
| Baseline metabolomic profiles: AC have prominent sugar metabolism whereas EC exhibit strong purine and amino acid metabolism | PMC7210983__F2 | pfocr | 0.010 | 5.8 |
| New insight into arginine and tryptophan metabolism in macrophage activation during tuberculosis | PMC11008464__F1 | pfocr | 0.010 | 14.3 |
| Major amino acid metabolic pathway involves AAD | PMC10411465__F3 | pfocr | 0.010 | 14.3 |
| Biosynthesis of AhR ligands from tryptophan metabolism | PMC6727512__F3 | pfocr | 0.011 | 13.3 |
| Sphingolipid metabolic pathway | PMC11185840__F2 | pfocr | 0.011 | 13.3 |
| Glycosphingolipid catabolism | R-HSA-9840310 | reactome | 0.012 | 7.9 |
| Visual representation generated by MetScape (CREGN) application of the Urea cycle and metabolism of arginine, proline, glutamate, aspartate and asparagine, a common pathway among the three species | PMC7598260__F7 | pfocr | 0.012 | 12.5 |
| Metabolic pathway of tryptophan (Trp) | PMC10943832__F2 | pfocr | 0.012 | 12.5 |
| Transcriptional response to a shift from aerobiosis via microaerobiosis to anaerobiosis including genes of glycolysis, tricarboxylic acid cycle (TCA), glyoxylate shunt, oxidative pentose phosphate pathway and selected amino acid biosynthesis pathways | PMC6027265__F3 | pfocr | 0.014 | 11.1 |
| Quantitative Metabolomics of Tissue, Perfusate, and Bile from Rat Livers Subjected to Normothermic Machine Perfusion | PMC8945564__F6 | pfocr | 0.016 | 10.0 |

| | | | | |
|---|---|---|---|---|
| Regulation mechanism diagram of DEGs and differential metabolites (fats and amino acids) | PMC10486348__F6 | pfocr | 0.016 | 10.0 |
| Metabolic pathways of the DFC involved in glucose lipid metabolism and amino acid metabolism | PMC10993571__F8 | pfocr | 0.018 | 9.5 |
| Amino Acid Biosynthesis Inhibitors in Tuberculosis Drug Discovery | PMC11206623__F20 | pfocr | 0.018 | 9.5 |
| GSH synthesis and transport of precursor amino acids: (a) Gamma-Glutamyl cycle of GSH synthesis | PMC11312684__F1 | pfocr | 0.018 | 9.5 |
| Lactobacillus plantarum and Acetobacter pasteurianus sequences mapped to a metabolic pathway for phenylalanine, tyrosine and tryptophan biosynthesis based on KEGG database | PMC6667825__F16 | pfocr | 0.020 | 8.7 |
| Summarized connections among the amino acid pathway affected by Lactobacillus reuteri (LR)in healthy breastfed mice | PMC6962498__F10 | pfocr | 0.028 | 7.1 |
| Amino acid metabolism pathway excerpt: histidine catabolism extension | WP4661 | wiki | 0.028 | 11.8 |
| Amino acid metabolism in triple-negative breast cancer cells | WP5213 | wiki | 0.035 | 10.5 |

Supplemental Figure 4. Metabolites significantly associated with OIT remission status, adjusted for age at enrollment. The coefficient represents the scaled change of metabolite concentration between OIT remission and non-remission.

| Metabolites | Subclass (RaMP) | HMDB | OR [95% CI] | p-value (GLM) |
|---|---|---|---|---|
| **Lysophospholipid** | | | | |
| 2-linoleoyl-GPC (18:2) | Glycerophosphocholines | HMDB0061700 | 0.26 [0.06, 0.99] | 0.048 |
| 1-oleoyl-GPE (18:1) | Glycerophosphoethanolamines | HMDB0011506 | 0.25 [0.07, 0.9] | 0.034 |
| 2-arachidonoyl-GPC (20:4) | Glycerophosphocholines | HMDB0061699 | 0.23 [0.06, 0.81] | 0.022 |
| 2-stearoyl-GPE (18:0) | Glycerophosphoethanolamines | HMDB0011129 | 0.22 [0.05, 0.85] | 0.027 |
| 1-palmitoleoyl-GPC (16:1) | Glycerophosphocholines | HMDB0010383 | 0.19 [0.04, 0.88] | 0.033 |
| 1-dihomo-linolenoyl-GPE (20:3n3 or 6) | Glycerophosphoethanolamines | HMDB0011514 | 0.18 [0.05, 0.69] | 0.011 |
| 1-linoleoyl-GPC (18:2) | Glycerophosphocholines | HMDB0010386 | 0.13 [0.02, 0.77] | 0.024 |
| 1-palmitoyl-GPE (16:0) | Glycerophosphoethanolamines | HMDB0011503 | 0.13 [0.02, 0.71] | 0.018 |
| 1-arachidonoyl-GPC (20:4n6) | Glycerophosphocholines | HMDB0010395 | 0.13 [0.02, 0.67] | 0.014 |
| 1-stearoyl-GPE (18:0) | Glycerophosphoethanolamines | HMDB0011130 | 0.12 [0.02, 0.66] | 0.014 |
| 1-arachidonoyl-GPE (20:4n6) | Glycerophosphoethanolamines | HMDB0011517 | 0.09 [0.02, 0.4] | 0.0016 |
| 1-oleoyl-GPC (18:1) | Glycerophosphocholines | HMDB0002815 | 0.04 [0, 0.38] | 0.004 |
| **Benzoate Metabolism** | | | | |
| 4-ethylphenyl sulfate | | HMDB0062551 | 9.63 [2.72, 47.91] | $3 \times 10^{-5}$ |
| 4-ethylcatechol sulfate | Glycosphingolipids | HMDB0004971 | 8.66 [2.6, 38.54] | $1.3 \times 10^{-4}$ |
| 3-hydroxyhippurate sulfate | | | 6.61 [1.82, 30.75] | 0.0027 |
| 4-allylcatechol sulfate | | | 5.93 [1.84, 24.27] | 0.0016 |
| 3-methoxycatechol sulfate (1) | Fatty acids and conjugates | HMDB0061659 | 4.71 [1.69, 15.9] | 0.0019 |
| 3-(3-hydroxyphenyl)propionate sulfate | Arylsulfates | HMDB0094710 | 3.97 [1.34, 13.89] | 0.012 |
| 4-acetylphenyl sulfate | | | 3.94 [1.37, 13.73] | 0.009 |
| 4-hydroxyhippurate | Benzoic acids and derivatives | HMDB0013678 | 3.61 [1.12, 13.18] | 0.031 |
| 4-vinylphenol sulfate | Arylsulfates | HMDB0062775 | 3.4 [1.28, 9.96] | 0.013 |
| 3-hydroxyhippurate | Benzoic acids and derivatives | HMDB0006116 | 3.05 [1.12, 9.25] | 0.028 |
| 3-(3-hydroxyphenyl)propionate | | HMDB0000375 | 2.59 [1.01, 7.38] | 0.048 |
| **Plasmalogen** | | | | |
| 1-(1-enyl-palmitoyl)-2-arachidonoyl-GPE (P-16:0/20:4) | Glycerophosphoethanolamines | HMDB0011352 | 0.26 [0.07, 0.94] | 0.039 |
| 1-(1-enyl-palmitoyl)-2-linoleoyl-GPE (P-16:0/18:2) | Glycerophosphoethanolamines | HMDB0011343 | 0.25 [0.07, 0.85] | 0.027 |
| 1-palmityl-2-linoleoyl-GPC (O-16:0/18:2) | Glycerophosphocholines | HMDB0011151 | 0.21 [0.04, 0.95] | 0.043 |
| 1-(1-enyl-stearoyl)-2-arachidonoyl-GPE (P-18:0/20:4) | Glycerophosphoethanolamines | HMDB0005779 | 0.18 [0.05, 0.62] | 0.0062 |
| 1-(1-enyl-stearoyl)-2-linoleoyl-GPE (P-18:0/18:2) | Glycerophosphoethanolamines | HMDB0011376 | 0.17 [0.04, 0.63] | 0.0077 |
| 1-(1-enyl-palmitoyl)-2-oleoyl-GPE (P-16:0/18:1) | Glycerophosphoethanolamines | HMDB0011342 | 0.16 [0.04, 0.62] | 0.0076 |
| 1-palmityl-2-arachidonoyl-GPC (O-16:0/20:4) | Glycerophosphocholines | HMDB0007982 | 0.13 [0.02, 0.72] | 0.018 |
| 1-(1-enyl-stearoyl)-2-linoleoyl-GPC (P-18:0/18:2) | Glycerophosphocholines | HMDB0011244 | 0.13 [0.02, 0.66] | 0.013 |
| 1-(1-enyl-stearoyl)-2-oleoyl-GPC (P-18:0/18:1) | Glycerophosphocholines | HMDB0011243 | 0.11 [0.02, 0.64] | 0.013 |
| 1-(1-enyl-stearoyl)-2-oleoyl-GPE (P-18:0/18:1) | Glycerophosphoethanolamines | HMDB0011375 | 0.11 [0.03, 0.39] | $5.9 \times 10^{-4}$ |
| 1-stearyl-2-linoleoyl-GPC (O-18:0/18:2) | Glycerophosphocholines | HMDB0013418 | 0.07 [0.01, 0.42] | 0.0033 |
| **Food Component/Plant** | | | | |
| 4-allylphenol sulfate | | HMDB0341325 | 8.05 [2.49, 32.57] | $2 \times 10^{-4}$ |
| 3-indoleglyoxylic acid | Medium-chain hydroxy acids and derivatives | HMDB0242119 | 6.85 [1.45, 39.25] | 0.014 |
| ferulic acid 4-sulfate | Hydroxycinnamic acids and derivatives | HMDB0029200 | 6.75 [2.17, 24.41] | $6.4 \times 10^{-4}$ |
| 4-acetylcatechol sulfate (1) | | | 4.92 [1.55, 17.99] | 0.0058 |
| caffeic acid sulfate | Hydroxycinnamic acids and derivatives | HMDB0041708 | 4.3 [1.48, 14.43] | 0.0064 |
| quinate | Alcohols and polyols | HMDB0003072 | 3.46 [1.44, 9.07] | 0.0049 |
| piperine | | HMDB0029377 | 0.5 [0.27, 0.89] | 0.018 |
| sulfate of piperine metabolite C16H19NO3 (3) | | | 0.48 [0.22, 1] | 0.049 |
| **Fatty Acid, Dicarboxylate** | | | | |
| dimethylmalonic acid | Dicarboxylic acids and derivatives | HMDB0002001 | 18.34 [3.25, 134.55] | $5.8 \times 10^{-4}$ |
| eicosanedioate (C20-DC) | Cinnamic acids | HMDB0125166 | 0.31 [0.09, 1] | 0.05 |
| dodecadienoate (12:2) | Fatty acids and conjugates | HMDB0245154 | 0.24 [0.07, 0.81] | 0.021 |
| 3-hydroxyadipate | Medium-chain hydroxy acids and derivatives | HMDB0000345 | 0.05 [0.01, 0.23] | $4.6 \times 10^{-5}$ |
| **Secondary Bile Acid Metabolism** | | | | |
| taurodeoxycholic acid 3-sulfate | | | 9.4 [2.6, 39.8] | $4 \times 10^{-4}$ |
| glycodeoxycholate 3-sulfate | Fatty acids and conjugates | HMDB0241923 | 8.31 [2.18, 38.11] | 0.0014 |
| taurocholenate sulfate | | | 5.97 [1.72, 23.65] | 0.0043 |
| taurolithocholate 3-sulfate | Bile acids, alcohols and derivatives | HMDB0002580 | 2.56 [1.01, 7.13] | 0.047 |
| **Chemical** | | | | |
| dimethyl sulfone | Sulfones | HMDB0004983 | 5.31 [1.63, 18.74] | 0.0054 |
| 3-acetylphenol sulfate | | | 4.91 [1.46, 19.55] | 0.0087 |
| perfluorooctanesulfonate (PFOS) | | HMDB0059586 | 3.54 [1.05, 13.64] | 0.04 |
| perfluorohexanesulfonate (PFHxS) | | HMDB0094700 | 2.98 [1.1, 9.44] | 0.031 |
| **Dipeptide** | | | | |
| cyclo(gly-pro) | Indolyl carboxylic acids and derivatives | HMDB0242143 | 5.07 [1.17, 28.46] | 0.028 |
| histidylalanine | Amino acids, peptides, and analogues | HMDB0028878 | 4.45 [1.18, 22.52] | 0.024 |
| glycylproline | Amino acids, peptides, and analogues | HMDB0000721 | 3.39 [1.13, 12.41] | 0.027 |
| leucylglycine | Amino acids, peptides, and analogues | HMDB0028929 | 3.31 [1.2, 11.24] | 0.018 |
| **Fatty Acid Metabolism (Acyl Carnitine, Medium Chain)** | | | | |
| cis-3,4-methyleneheptanoylcarnitine | | | 0.34 [0.11, 0.98] | 0.045 |
| laurylcarnitine (C12) | Medium-chain keto acids and derivatives | HMDB0000225 | 0.29 [0.1, 0.86] | 0.026 |
| decanoylcarnitine (C10) | Fatty acid esters | HMDB0000651 | 0.15 [0.05, 0.41] | $2.1 \times 10^{-4}$ |
| octanoylcarnitine (C8) | Fatty acid esters | HMDB0000791 | 0.1 [0.03, 0.3] | $2.5 \times 10^{-5}$ |
| **Fatty Acid Metabolism (Acyl Carnitine, Monounsaturated)** | | | | |
| 5-dodecenoylcarnitine (C12:1) | Fatty acid esters | HMDB0013326 | 0.27 [0.09, 0.79] | 0.017 |
| myristoleoylcarnitine (C14:1) | Fatty acid esters | HMDB0240588 | 0.19 [0.06, 0.62] | 0.0057 |
| cis-4-decenoylcarnitine (C10:1) | Fatty acid esters | HMDB0013205 | 0.15 [0.04, 0.47] | $9.4 \times 10^{-4}$ |
| ximenoylcarnitine (C26:1) | | | 0.13 [0.02, 0.76] | 0.024 |
| **Phosphatidylethanolamine (PE)** | | | | |
| 1-palmitoyl-2-arachidonoyl-GPE (16:0/20:4) | | HMDB0005323 | 0.26 [0.07, 0.91] | 0.035 |
| 1-stearoyl-2-dihomo-linolenoyl-GPE (18:0/20:3n3 or 6) | Glycerophosphoethanolamines | HMDB0009002 | 0.25 [0.07, 0.81] | 0.02 |
| 1-palmitoyl-2-docosahexaenoyl-GPE (16:0/22:6) | Glycerophosphoethanolamines | HMDB0008946 | 0.22 [0.06, 0.79] | 0.02 |
| 1-stearoyl-2-arachidonoyl-GPE (18:0/20:4) | Glycerophosphoethanolamines | HMDB0009003 | 0.16 [0.04, 0.58] | 0.0049 |
| **Tryptophan Metabolism** | | | | |
| kynurenate | Quinoline carboxylic acids | HMDB0000715 | 8.38 [1.68, 49.8] | 0.0088 |
| indolelactate | Indolyl carboxylic acids and derivatives | HMDB0000671 | 6.67 [1.26, 40.7] | 0.025 |
| 5-hydroxyindoleacetate | Indolyl carboxylic acids and derivatives | HMDB0000763 | 6.47 [1.41, 34.86] | 0.015 |
| **Primary Bile Acid Metabolism** | | | | |
| glycochenodeoxycholate glucuronide (1) | Steroidal glycosides | HMDB0002579 | 2.89 [1.08, 8.62] | 0.035 |
| glyco-beta-muricholate | | | 2.87 [1.14, 8.27] | 0.024 |
| tauro-beta-muricholate | Bile acids, alcohols and derivatives | HMDB0000932 | 2.83 [1.17, 7.71] | 0.02 |
| **Androgenic Steroids** | | | | |
| androsterone sulfate | Sulfated steroids | HMDB0002759 | 0.32 [0.14, 0.72] | 0.0059 |
| dehydroepiandrosterone sulfate (DHEA-S) | Sulfated steroids | HMDB0001032 | 0.31 [0.14, 0.69] | 0.0037 |
| androstenediol (3beta,17beta) disulfate (1) | Sulfated steroids | HMDB0240313 | 0.28 [0.11, 0.7] | 0.0059 |
| **Fatty Acid Metabolism (Acyl Carnitine, Long Chain Saturated)** | | | | |
| stearoylcarnitine (C18) | Fatty acid esters | HMDB0000848 | 0.3 [0.09, 1] | 0.049 |
| lignoceroylcarnitine (C24) | Fatty acid esters | HMDB0240665 | 0.15 [0.03, 0.75] | 0.021 |
| arachidoylcarnitine (C20) | Fatty acid esters | HMDB0006460 | 0.07 [0.01, 0.35] | 0.001 |
| **Tyrosine Metabolism** | | | | |
| tyrosine | Amino acids, peptides, and analogues | HMDB0000158 | 8.42 [1.6, 52.08] | 0.011 |
| 3-methoxytyrosine | Amino acids, peptides, and analogues | HMDB0001434 | 0.13 [0.02, 0.77] | 0.024 |
| **Leucine, Isoleucine and Valine Metabolism** | | | | |
| valine | Amino acids, peptides, and analogues | HMDB0000883 | 6.72 [1.18, 45.6] | 0.032 |
| isovalerate (C5) | Fatty acids and conjugates | HMDB0000718 | 4.08 [1, 19.31] | 0.05 |
| **Urea cycle; Arginine and Proline Metabolism** | | | | |
| arginine | Amino acids, peptides, and analogues | HMDB0000517 | 6.53 [1.17, 44.81] | 0.032 |
| N,N,N-trimethyl-alanylproline betaine (TMAP) | Amino acids, peptides, and analogues | HMDB0240365 | 0.02 [0, 0.14] | $1.1 \times 10^{-4}$ |
| **Lysine Metabolism** | | | | |
| 6-oxopiperidine-2-carboxylate | Amino acids, peptides, and analogues | HMDB0061705 | 4.73 [1.18, 22.26] | 0.027 |
| N6-methyllysine | Amino acids, peptides, and analogues | HMDB0002038 | 4.16 [1.32, 16.22] | 0.013 |
| **Drug - Topical Agents** | | | | |
| hydroquinone sulfate | Arylsulfates | HMDB0240263 | 4.25 [1.29, 17.52] | 0.015 |
| 2,6-dihydroxybenzoic acid | Benzoic acids and derivatives | HMDB0013676 | 3.64 [1.12, 13.28] | 0.031 |
| **Partially Characterized Molecules** | | | | |
| glutamine conjugate of C7H12O2 | | | 0.39 [0.15, 0.95] | 0.038 |
| glutamine conjugate of C6H10O2 (2) | | | 0.33 [0.11, 0.92] | 0.034 |
| **Phosphatidylcholine (PC)** | | | | |
| phosphatidylcholine (18:0/20:2, 20:0/18:2) | Glycerophosphocholines | HMDB0008143 | 0.18 [0.04, 0.85] | 0.03 |
| 1-stearoyl-2-arachidonoyl-GPC (18:0/20:4) | Glycerophosphocholines | HMDB0008048 | 0.17 [0.03, 0.97] | 0.046 |
| **Creatine Metabolism** | | | | |
| guanidinoacetate | Amino acids, peptides, and analogues | HMDB0000128 | 0.18 [0.03, 0.94] | 0.042 |
| creatinine | Amino acids, peptides, and analogues | HMDB0000562 | 0.11 [0.01, 0.81] | 0.03 |
| **Hexosylceramides (HCER)** | | | | |
| glycosyl ceramide (d18:2/24:1, d18:1/24:2) | | | 0.16 [0.04, 0.62] | 0.0079 |
| glycosyl-N-behenoyl-sphingosine (d18:1/22:0) | | | 0.15 [0.03, 0.7] | 0.015 |
| **Sphingomyelins** | | | | |
| sphingomyelin (d18:2/18:1) | Phosphosphingolipids | HMDB0001348 | 0.09 [0.01, 0.71] | 0.021 |
| sphingomyelin (d18:2/16:0, d18:1/16:1) | Phosphosphingolipids | HMDB0240638 | 0.08 [0.01, 0.8] | 0.031 |
| **Guanidino and Acetamido Metabolism** | | | | |
| 4-guanidinobutanoate | Amino acids, peptides, and analogues | HMDB0003464 | 7.03 [1.45, 40.05] | 0.014 |
| **Bacterial/Fungal** | | | | |
| N-methylpipecolate | Fatty amides | HMDB0242113 | 5.48 [1.49, 24.79] | 0.0092 |
| **Fibrinogen Cleavage Peptide** | | | | |
| fibrinopeptide B (1-11) | Arylsulfates | HMDB0242163 | 3.45 [1.03, 15.59] | 0.044 |
| **Fructose, Mannose and Galactose Metabolism** | | | | |
| galactonate | Medium-chain hydroxy acids and derivatives | HMDB0000565 | 3.25 [1.22, 9.5] | 0.018 |
| **Fatty Acid, Dihydroxy** | | | | |
| 9,10-DiHOME | Fatty acids and conjugates | HMDB0004704 | 2.84 [1.03, 8.95] | 0.044 |
| **Fatty Acid Metabolism (Acyl Carnitine, Short Chain)** | | | | |
| acetylcarnitine (C2) | Fatty acid esters | HMDB0000201 | 0.45 [0.2, 0.95] | 0.037 |
| **Endocannabinoid** | | | | |
| N-stearoyltaurine | Indolyl carboxylic acids and derivatives | HMDB0242168 | 0.31 [0.1, 0.96] | 0.042 |
| **Fatty Acid Metabolism (Acyl Glycine)** | | | | |
| N-palmitoylglycine | Amino acids, peptides, and analogues | HMDB0013034 | 0.29 [0.1, 0.83] | 0.02 |
| **Fatty Acid, Monohydroxy** | | | | |
| 3-hydroxydecanoate | Medium-chain hydroxy acids and derivatives | HMDB0002203 | 0.27 [0.07, 0.93] | 0.039 |
| **Histidine Metabolism** | | | | |
| imidazole lactate | Amino acids (histidine metabolism) | HMDB0002320 | 0.25 [0.06, 0.99] | 0.048 |
| **Medium Chain Fatty Acid** | | | | |
| cis-4-decenoate (10:1n6) | Fatty acids and conjugates | HMDB0004980 | 0.23 [0.07, 0.75] | 0.015 |
| **Pyrimidine Metabolism, Uracil containing** | | | | |
| N-acetyl-beta-alanine | Carboxylic acids | HMDB0061880 | 0.22 [0.05, 0.96] | 0.043 |
| **Fatty Acid Metabolism (Acyl Carnitine, Dicarboxylate)** | | | | |
| adipoylcarnitine (C6-DC) | Fatty acid esters | HMDB0061677 | 0.22 [0.07, 0.69] | 0.0086 |
| **Polyamine Metabolism** | | | | |
| N-acetyl-isoputreanine | Amino acids, peptides, and analogues | HMDB0242185 | 0.2 [0.04, 1] | 0.05 |
| **Sterol** | | | | |
| 3beta,7beta-dihydroxy-5-cholestenoate | | | 0.17 [0.03, 0.78] | 0.022 |
| **Methionine, Cysteine, SAM and Taurine Metabolism** | | | | |
| succinoyltaurine | | | 0.08 [0.01, 0.34] | $5.1 \times 10^{-4}$ |
| **Glutamate Metabolism** | | | | |
| N-acetylglutamate | Amino acids, peptides, and analogues | HMDB0001138 | 0.06 [0.01, 0.25] | $5.7 \times 10^{-5}$ |

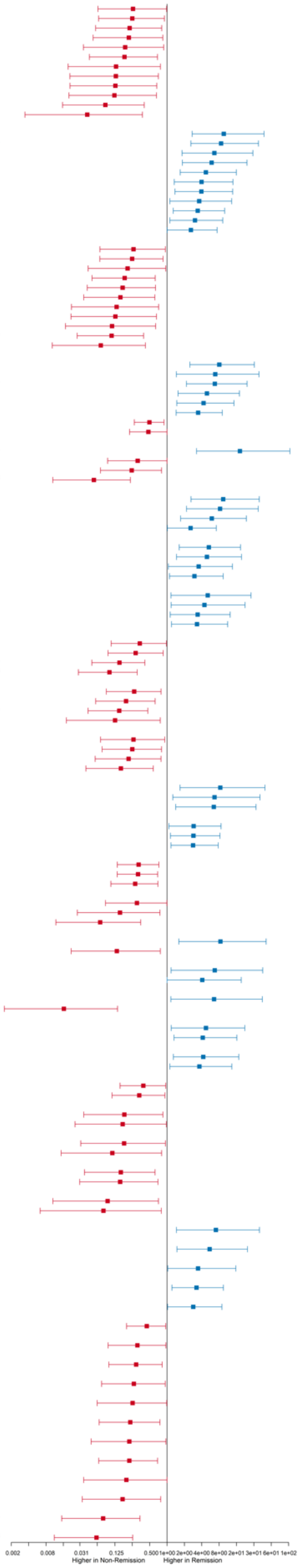

Supplemental Figure 5. Biological pathways enrichment of metabolites associated with remission status by RAMP-based biological pathway analysis. The percent of matched metabolites in pathway plotted by the FDR-adjusted enrichment p-value. Dark pink represents pathways linked to food allergy in prior publications.

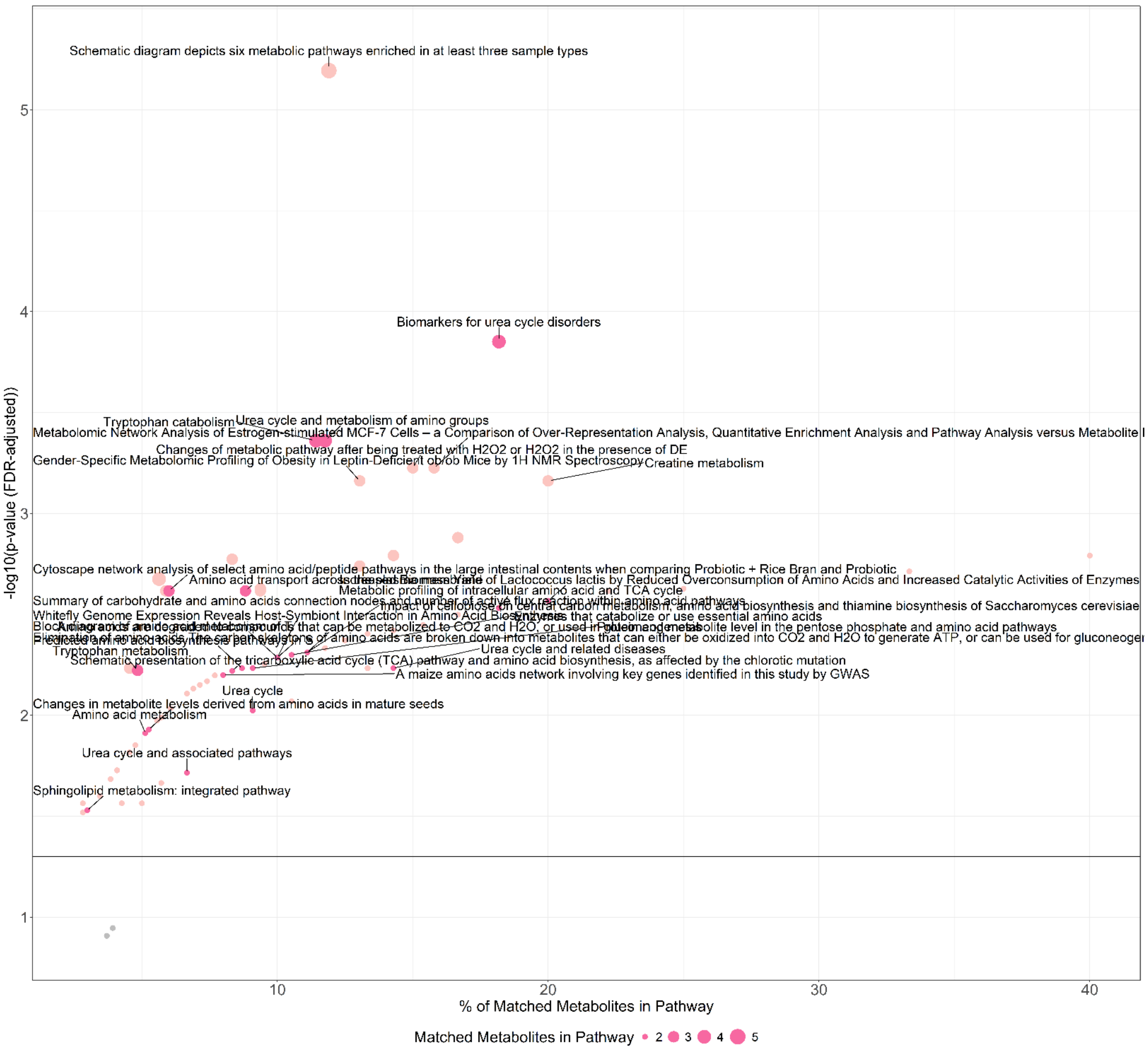

Supplemental Figure 6. Biological pathways significantly associated with remission status. Pathways enriched in both *DEVIL* and *PNOIT* are indicated in orange and scaled by the percent of matching metabolites.

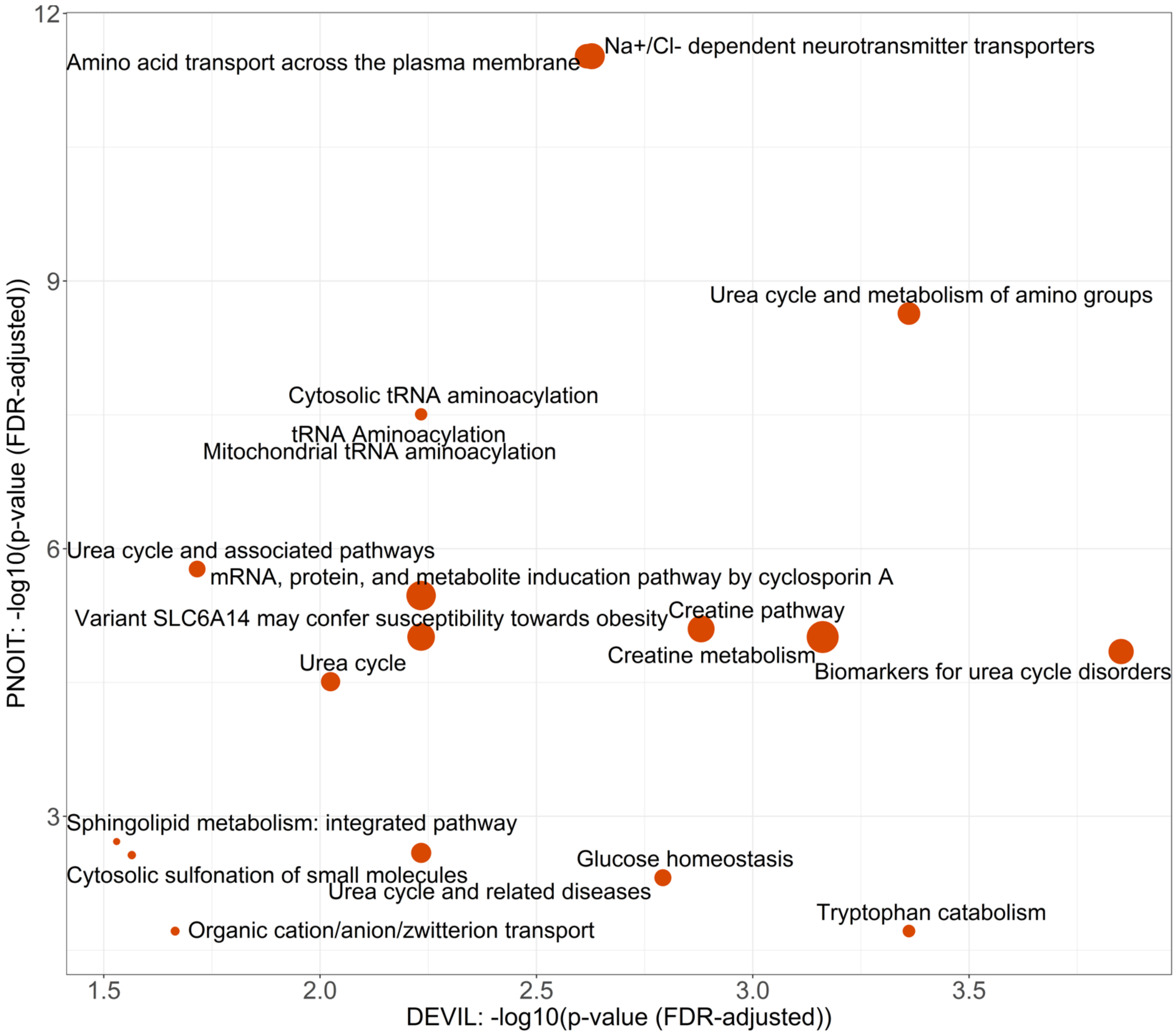

Supplemental Table 6. Summary of chemical subclass enrichment of metabolites significantly associated with OIT remission status.

| Subclass | P-val (FDR adjusted) | Enrichment Ratio | Percent Hits (%) |
|---|---|---|---|
| Glycerophosphocholines | 4.30E-10 | 13.6 | 1.0 |
| Glycerophosphoethanolamines | 4.28E-08 | 6.9 | 0.5 |
| Amino acids, peptides, and analogues | 4.28E-08 | 6.3 | 0.4 |
| Arylsulfates | 9.30E-08 | 67.1 | 4.7 |
| Indolyl carboxylic acids and derivatives | 5.44E-07 | 94.8 | 6.7 |
| Medium-chain hydroxy acids and derivatives | 7.10E-07 | 84.9 | 6.0 |
| Fatty acid esters | 1.16E-06 | 8.5 | 0.6 |
| Sulfated steroids | 1.78E-05 | 88.9 | 6.3 |
| Bile acids, alcohols and derivatives | 0.00014 | 20.2 | 1.4 |
| Fatty acids and conjugates | 0.00027 | 8.1 | 0.6 |
| Phosphosphingolipids | 0.00447 | 30.9 | 2.2 |
| Cinnamic acids | 0.015 | 142.2 | 10.0 |
| Sulfones | 0.015 | 129.3 | 9.1 |
| Benzoic acids and derivatives | 0.021 | 6.5 | 0.5 |
| Hydroxycinnamic acids and derivatives | 0.023 | 11.4 | 0.8 |
| Carboxylic acids | 0.043 | 35.5 | 2.5 |
| Medium-chain keto acids and derivatives | 0.046 | 31.6 | 2.2 |
| Fatty amides | 0.063 | 21.5 | 1.5 |
| Quinoline carboxylic acids | 0.066 | 19.5 | 1.4 |
| Dicarboxylic acids and derivatives | 0.082 | 14.8 | 1.0 |
| Imidazoles | 0.11 | 10.4 | 0.7 |
| Steroidal glycosides | 0.20 | 5.2 | 0.4 |
| Alcohols and polyols | 0.23 | 4.1 | 0.3 |
| Glycosphingolipids | 0.31 | 2.8 | 0.2 |
| Carbonyl compounds | 0.52 | 1.4 | 0.1 |

Supplemental Table 7. Summary of biological pathways enriched based on metabolites significantly associated with OIT remission status.

| Biological Pathway | Pathway ID | Pathway Source | P-val (FDR adjusted) | % Matched Metabolites |
|---|---|---|---|---|
| Biomarkers for urea cycle disorders | WP4583 | wiki | 0.0001 | 18.2 |
| Tryptophan catabolism | R-HSA-71240 | reactome | 0.0004 | 11.4 |
| Urea cycle and metabolism of amino groups | WP497 | wiki | 0.0004 | 11.8 |
| Amino acid transport across the plasma membrane | R-HSA-352230 | reactome | 0.0024 | 8.8 |
| Cytoscape network analysis of select amino acid/peptide pathways in the large intestinal contents when comparing Probiotic + Rice Bran and Probiotic | PMC5399067__F5 | pfocr | 0.0024 | 6.0 |
| Increased Biomass Yield of Lactococcus lactis by Reduced Overconsumption of Amino Acids and Increased Catalytic Activities of Enzymes | PMC3485057__F4 | pfocr | 0.0027 | 20.0 |
| Summary of carbohydrate and amino acids connection nodes and number of active flux reaction within amino acid pathways | PMC4236713__F6 | pfocr | 0.0029 | 18.2 |
| Metabolic profiling of intracellular amino acid and TCA cycle | PMC5501892__F3 | pfocr | 0.0029 | 18.2 |
| Amino acids are degraded to compounds that can be metabolized to CO2 and H2O, or used in gluconeogenesis | PMC3156598__F1 | pfocr | 0.0049 | 11.1 |
| Impact of cellobiose on central carbon metabolism, amino acid biosynthesis and thiamine biosynthesis of Saccharomyces cerevisiae | PMC4243952__F2 | pfocr | 0.0049 | 11.1 |
| Enzymes that catabolize or use essential amino acids | PMC3839643__F3 | pfocr | 0.0050 | 10.5 |
| Block diagram of amino acid metabolism of T | PMC16270__F1 | pfocr | 0.0052 | 10.0 |
| Whitefly Genome Expression Reveals Host-Symbiont Interaction in Amino Acid Biosynthesis | PMC4441466__F7 | pfocr | 0.0052 | 10.0 |
| Elimination of amino-acids.The carbon skeletons of amino acids are broken down into metabolites that can either be oxidized into CO2 and H2O to generate ATP, or can be used for gluconeogenesis | PMC5551501__F2 | pfocr | 0.0052 | 10.0 |
| Urea cycle and related diseases | WP4571 | wiki | 0.0058 | 14.3 |
| Predicted amino acid biosynthesis pathways in S | PMC2739083__F5 | pfocr | 0.0058 | 8.7 |
| Protein and metabolite level in the pentose phosphate and amino acid pathways | PMC4404605__F4 | pfocr | 0.0058 | 9.1 |
| Tryptophan metabolism | WP465 | wiki | 0.0060 | 4.8 |
| Schematic presentation of the tricarboxylic acid cycle (TCA) pathway and amino acid biosynthesis, as affected by the chlorotic mutation | PMC5337497__F3 | pfocr | 0.0060 | 8.3 |
| A maize amino acids network involving key genes identified in this study by GWAS | PMC5595712__F3 | pfocr | 0.0063 | 8.0 |
| Urea cycle | R-HSA-70635 | reactome | 0.0095 | 9.1 |
| Changes in metabolite levels derived from amino acids in mature seeds | PMC3901229__F4 | pfocr | 0.012 | 5.3 |
| Amino acid metabolism | PMC10497558__F1 | pfocr | 0.012 | 5.1 |

| | | | | |
|---|---|---|---|---|
| Urea cycle and associated pathways | WP4595 | wiki | 0.019 | 6.7 |
| Sphingolipid metabolism: integrated pathway | WP4726 | wiki | 0.030 | 3.0 |

Supplemental Table 8. Summary of chemical subclass enrichment of metabolites significantly associated with OIT remission status in both *DEVIL* and *PNOIT*. FDR corrected p-values are Benjamini-Hochberg corrected.

| **Subclass** | **P-val (FDR adjusted) (*DEVIL*)** | **Enrichment Ratio (*DEVIL*)** | **P-val (FDR adjusted) (*PNOIT*)** | **Enrichment Ratio (*PNOIT*)** | **Enrichment Ratio Mean (*DEVIL*& *PNOIT)*** |
|---|---|---|---|---|---|
| Glycerophosphocholines | 4.2988E-10 | 13.6 | 5.7018E-16 | 17.8 | 15.7 |
| Amino acids, peptides, and analogues | 4.284E-08 | 6.3 | 7.2249E-07 | 5.6 | 5.9 |
| Glycerophosphoethanolamines | 4.284E-08 | 6.9 | 0.0093 | 3.5 | 5.2 |
| Arylsulfates | 9.3008E-08 | 67.1 | | | 67.1 |
| Indolyl carboxylic acids and derivatives | 5.4444E-07 | 94.8 | 0.076 | 22.3 | 58.6 |
| Medium-chain hydroxy acids and derivatives | 7.1046E-07 | 84.9 | 0.080 | 20.0 | 52.4 |
| Fatty acid esters | 1.1571E-06 | 8.5 | 0.76 | 0.8 | 4.7 |
| Sulfated steroids | 1.7825E-05 | 88.9 | | | 88.9 |
| Bile acids, alcohols and derivatives | 0.00014 | 20.2 | 3.8172E-08 | 33.3 | 26.7 |
| Fatty acids and conjugates | 0.00027 | 8.1 | 7.7073E-07 | 11.4 | 9.7 |
| Phosphosphingolipids | 0.00447 | 30.9 | 0.096 | 14.6 | 22.7 |
| Cinnamic acids | 0.015 | 142.2 | | | 142.2 |
| Sulfones | 0.015 | 129.3 | | | 129.3 |
| Benzoic acids and derivatives | 0.021 | 6.5 | 0.44 | 2.0 | 4.2 |
| Hydroxycinnamic acids and derivatives | 0.023 | 11.4 | | | 11.4 |
| Carboxylic acids | 0.043 | 35.5 | | | 35.5 |
| Medium-chain keto acids and derivatives | 0.046 | 31.6 | | | 31.6 |
| Fatty amides | 0.063 | 21.5 | | | 21.5 |
| Quinoline carboxylic acids | 0.066 | 19.5 | | | 19.5 |
| Dicarboxylic acids and derivatives | 0.082 | 14.8 | 0.096 | 14.0 | 14.4 |
| Imidazoles | 0.11 | 10.4 | 0.12 | 9.8 | 10.1 |
| Steroidal glycosides | 0.20 | 5.2 | | | 5.2 |
| Alcohols and polyols | 0.23 | 4.1 | | | 4.1 |
| Glycosphingolipids | 0.31 | 2.8 | | | 2.8 |
| Carbonyl compounds | 0.52 | 1.4 | 0.59 | 1.3 | 1.3 |

| | | | | | |
|---|---|---|---|---|---|
| Eicosanoids | | | 7.2249E-07 | 19.5 | 19.5 |
| Purine ribonucleotides | | | 4.087E-06 | 58.9 | 58.9 |
| Quaternary ammonium salts | | | 0.0039 | 47.9 | 47.9 |
| Sulfinic acids | | | 0.014 | 268.1 | 268.1 |
| Alpha hydroxy acids and derivatives | | | 0.020 | 167.6 | 167.6 |
| Steroid esters | | | 0.029 | 13.6 | 13.6 |
| Pyrrolidinylpyridines | | | 0.037 | 74.5 | 74.5 |
| Pyrimidines and pyrimidine derivatives | | | 0.038 | 10.6 | 10.6 |
| Gamma-keto acids and derivatives | | | 0.038 | 58.3 | 58.3 |
| Purines and purine derivatives | | | 0.038 | 10.0 | 10.0 |
| Guanidines | | | 0.061 | 33.5 | 33.5 |
| Amines | | | 0.063 | 7.1 | 7.1 |
| Organosulfonic acids and derivatives | | | 0.076 | 23.1 | 23.1 |
| Tricarboxylic acids and derivatives | | | 0.096 | 15.6 | 15.6 |
| Pyridinecarboxylic acids and derivatives | | | 0.096 | 13.8 | 13.8 |
| Monoradylglycerols | | | 0.11 | 11.8 | 11.8 |
| Linoleic acids and derivatives | | | 0.29 | 3.7 | 3.7 |
| Carbohydrates and carbohydrate conjugates | | | 0.44 | 1.5 | 1.5 |
| Diradylglycerols | | | 0.91 | 0.5 | 0.5 |
| Triacylglycerols | | | 1.0 | 0.2 | 0.2 |

Supplemental Table 9. Summary of biological pathways enriched based on metabolites significantly associated with OIT remission status in both *DEVIL* and *PNOIT*.

| **Biological Pathway** | **Pathway ID** | **Pathway Source** | **P-val (FDR adjusted) (DEVIL)** | **% Matched Metabolites (*DEVIL*)** | **P-val (FDR adjusted) (PNOIT)** | **% Matched Metaboites (*PNOIT*)** |
|---|---|---|---|---|---|---|
| Na+/Cl- dependent neurotransmitter transporters | R-HSA-442660 | reactome | 0.002 | 9.4 | 3.01E-12 | 28.1 |
| Amino acid transport across the plasma membrane | R-HSA-352230 | reactome | 0.002 | 8.8 | 3.01E-12 | 26.5 |
| Urea cycle and metabolism of amino groups | WP497 | wiki | 0.0004 | 11.8 | 2.32E-09 | 20.6 |
| Cytosolic tRNA aminoacylation | R-HSA-379716 | reactome | 0.006 | 4.5 | 3.12E-08 | 12.1 |
| tRNA Aminoacylation | R-HSA-379724 | reactome | 0.006 | 4.5 | 3.12E-08 | 12.1 |
| Mitochondrial tRNA aminoacylation | R-HSA-379726 | reactome | 0.006 | 4.5 | 3.12E-08 | 12.1 |
| Arginine and proline metabolism | map00330 | kegg | 0.11 | 3.9 | 6.21E-07 | 19.6 |

| | | | | | | |
|---|---|---|---|---|---|---|
| Urea cycle and associated pathways | WP4595 | wiki | 0.019 | 6.7 | 1.69E-06 | 16.7 |
| mRNA, protein, and metabolite inducation pathway by cyclosporin A | WP3953 | wiki | 0.006 | 14.3 | 3.35E-06 | 28.6 |
| Creatine pathway | WP5190 | wiki | 0.001 | 16.7 | 7.92E-06 | 22.2 |
| Creatine metabolism | R-HSA-71288 | reactome | 0.0007 | 20.0 | 9.76E-06 | 26.7 |
| Variant SLC6A14 may confer susceptibility towards obesity | R-HSA-5619094 | reactome | 0.006 | 13.3 | 9.76E-06 | 26.7 |
| Biomarkers for urea cycle disorders | WP4583 | wiki | 0.0001 | 18.2 | 1.42E-05 | 18.2 |
| Urea cycle | R-HSA-70635 | reactome | 0.009 | 9.1 | 3.08E-05 | 18.2 |
| Sphingolipid metabolism: integrated pathway | WP4726 | wiki | 0.030 | 3.0 | 0.0019 | 5.9 |
| Urea cycle and related diseases | WP4571 | wiki | 0.006 | 14.3 | 0.0026 | 14.3 |
| Cytosolic sulfonation of small molecules | R-HSA-156584 | reactome | 0.027 | 4.3 | 0.0027 | 6.4 |
| Glucose homeostasis | WP661 | wiki | 0.002 | 14.3 | 0.0049 | 9.5 |
| Tryptophan catabolism | R-HSA-71240 | reactome | 0.0004 | 11.4 | 0.019 | 5.7 |
| Organic cation/anion/zwitterion transport | R-HSA-549132 | reactome | 0.022 | 5.7 | 0.019 | 5.7 |
| Tryptophan metabolism | map00380 | kegg | 0.12 | 3.7 | 0.028 | 7.4 |
